# Childhood vaccination coverage by socioeconomic, geographic, maternal, child, and healthcare characteristics in Nigeria: equity impact analysis

**DOI:** 10.1101/2022.01.24.22269771

**Authors:** Sarah V Williams, Tanimola Akande, Kaja Abbas

**Affiliations:** Royal Free London, NHS Foundation Trust, NHS Foundation Trust, Pond Street, London, NW3 2QG, United Kingdom; Department of Epidemiology & Community Health, University of Ilorin, Nigeria; Department of Infectious Disease Epidemiology, London School of Hygiene & Tropical Medicine, Keppel Street, London, WC1E 7HT United Kingdom

**Keywords:** Childhood immunisation, vaccine equity, social determinants of immunisation, demographic and health survey, Nigeria

## Abstract

**Background:** Nigeria has a high proportion of the world’s underimmunised children. We estimated the inequities in childhood immunisation coverage associated with socioeconomic, geographic, maternal, child, and healthcare characteristics among children aged 12-23 months in Nigeria using a social determinants of health perspective.

**Methods:** We first conducted a systematic review focused on inequities in childhood immunisation in low- and middle-income countries to identify the social determinants of childhood immunisation. To identify inequities in basic vaccination coverage in Nigeria (1-dose BCG, 3-dose DTP-HepB-Hib (diphtheria, tetanus, pertussis, hepatitis B and Haemophilus influenzae type B), 3-dose polio, and 1-dose measles) of 6,059 children aged 12-23 months using the 2018 Nigeria Demographic and Health Survey, we conducted multiple logistic regression to analyse the association of basic vaccination with these determinants.

**Results:** We identified social determinants of immunisation: household wealth, religion, and ethnicity (socioeconomic characteristics); region and place of residence (geographic characteristics); maternal age at birth, maternal education, and maternal household head status (maternal characteristics); sex of child, and birth order (child characteristics); antenatal care and birth setting (healthcare characteristics). While basic vaccination coverage was 31% (95% CI: 29–33), 19% (95% CI:18–21) of children aged 12-23 months had received none of the basic vaccines. After controlling for background characteristics, there was a significant increase in the odds of basic vaccination by household wealth, antenatal care, delivery in a health facility, maternal age and maternal education. Children of Fulani ethnicity in comparison to children of Igbo ethnicity had lower odds of receiving basic vaccinations.

**Conclusions:** Children from the poorest households, of Fulani ethnicity, who were born in home settings, and with young mothers with no formal education nor antenatal care, were associated with lower odds of basic vaccination in Nigeria. Basic vaccination coverage is below target levels for all groups and we recommend a proportionate universalism approach for addressing the immunisation barriers.

## Introduction

Nigeria is the most populous country in Africa with around 202 million people in 2020 and its population is predicted to double by 2050 [1]. It is a multi-ethnic country with 36 autonomous states and the Federal Capital Territory. Around 83 million people (40% of total population) live below the poverty line while an additional 53 million people (25% of total population) are vulnerable to falling below the poverty line [2]. Economic growth has been slow with challenges including ongoing conflict in parts of the country, inconsistent regulatory environment, poor power supply and infrastructure [2].

Vaccination is a highly cost-effective public health intervention and beyond the direct benefits to population health, vaccines provide additional economic and social benefits to individuals and society [3, 4]. Infectious diseases remain a leading cause of death among under-5-year-old children, and an additional 1.5 million deaths could be avoided every year with improvements in global vaccination coverage [5, 6]. Model-based estimates, not including COVID-19, project 51 million deaths to be prevented by vaccination during 2021-2030 [7]. There have been substantial improvements in vaccine introductions and vaccination coverage in low- and middle-income countries since the inception of Gavi, the Vaccine Alliance in 2000 [8]. However, the prevalence of zero-dose children, that is children aged 12-23 months who had not received any of the routine childhood vaccines, was 7.7% in low- and middle-income countries during 2010-2019 [9]. The importance of improving vaccination coverage was recognised in the Sustainable Development Goals (SDGs), with immunisation contributing to 14 of the 17 SDGs and includes reduction on poverty and hunger and improving social equity [10]. Vaccination coverage and equity are a strategic goal of the global Immunisation Agenda 2030, with the aim to reach equitable coverage at national and district levels by addressing immunisation barriers posed by location, age, socioeconomic status, and gender [11].

The Expanded Programme on Immunisation (EPI) was established by the World Health Organization (WHO) in 1974 to improve vaccination services globally [12], and Nigeria began nationwide implementation of EPI in 1979, [13] which was later changed to the National Programme on Immunization. Although the vaccines in the routine immunisation programme (Table A1) for under 5-year-old children are available with no out-of-pocket charges [14], Nigeria has the most under-immunised children in the world with 4.5 million in 2018 [15]. The immunisation system challenges in Nigeria include weak institutions, service delivery, funding, infrastructure, poor coordination between the National Programme on Immunization and non-governmental organisations delivering vaccination services, and a lack of political commitment in some regions [14, 16]. There are fewer adequately skilled healthcare personnel in rural areas and northern states, and poor retention and frequent transfers of workers. Security is also an issue, with attacks on healthcare workers in recent years. Attitudes of communities and caregivers are important too, with a lack of knowledge about vaccination and mistrust of services hindering vaccination uptake [16, 17].

In the context of wider immunisation system challenges in Nigeria, we focused on factors associated with inequities in basic vaccination coverage through the social determinants of health model. This model framework has been explicitly linked to health equity by the WHO Commission on Social Determinants of Health [18] and considers the social, cultural, political, economic, commercial and environmental factors that shape the conditions in which people are born, grow, live, work and age, and these factors are determined by wealth, power and resources. In this study, we use the social determinants of health model framework, which encompasses the individual, parental, household, environment, and national policy levels that influence inequities in basic vaccination coverage among children in Nigeria (see Figure 1). We refer to vaccine inequity as unfair and avoidable or remediable differences in health among population groups defined socially, economically, demographically, or geographically [19]. This is related to but distinct from health inequality, which indicates the status of imbalances or differences in health among population groups without any moral judgement on whether the imbalances or differences are fair or not [20, 21].

Our aim is to analyse the 2018 Nigeria Demographic and Health Survey (DHS) and estimate the inequities in basic vaccination coverage (1-dose BCG, 3-dose DTP-HepB-Hib, 3-dose polio, and 1-dose measles vaccines) associated with socioeconomic, geographic, maternal, child, and healthcare characteristics among children aged 12-23 months in Nigeria.

**Figure 1.**
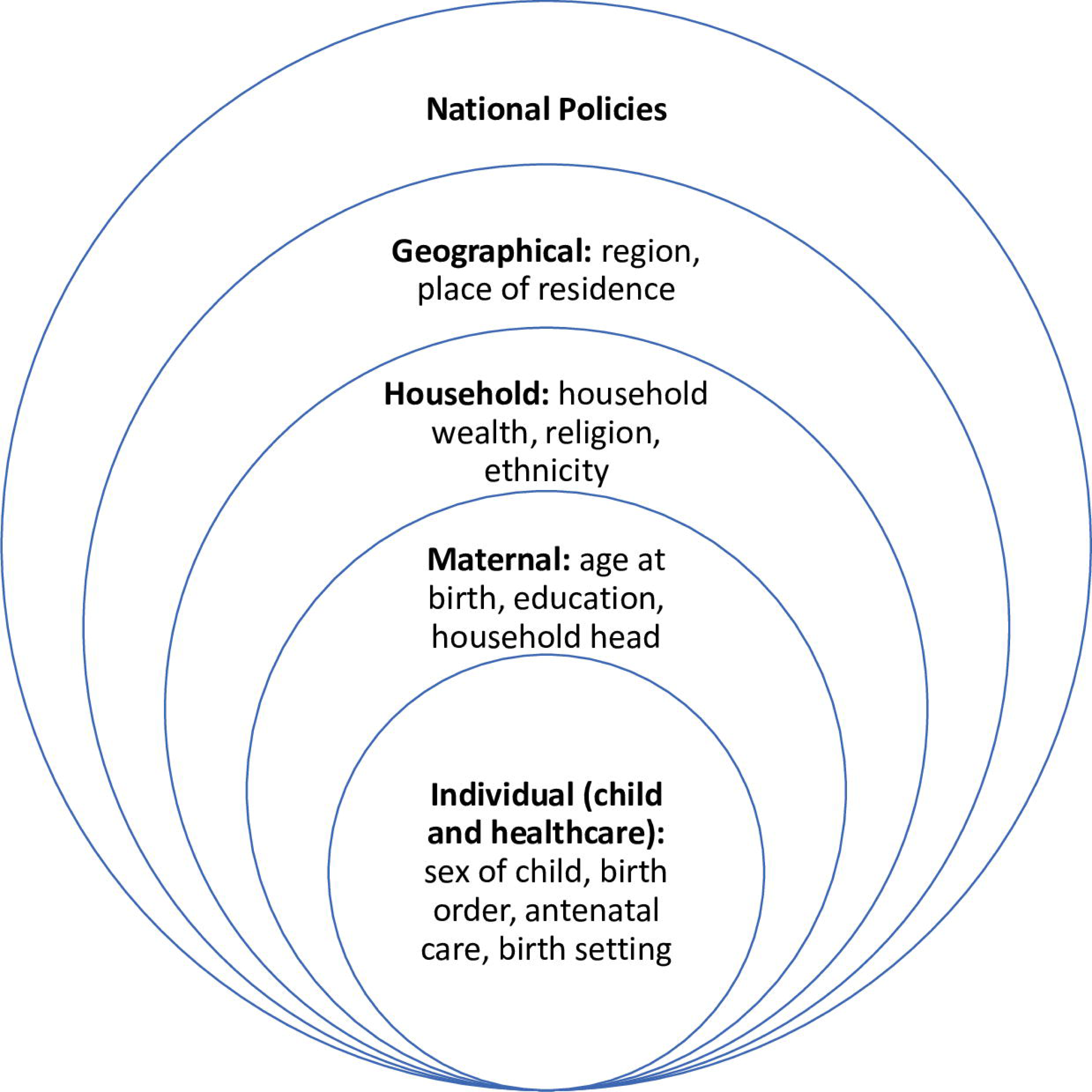
Social determinants of childhood immunisation. Social determinants of health model framework encompassing the individual, parental, household, environment, and national policy levels and influencing inequities in basic vaccination coverage among children in Nigeria.

## Methods

### Characteristics selection and systematic review

The WHO had analysed inequality in immunisation using the following characteristics determined from a literature review: child (gender, birth order), maternal (age at birth, education, ethnicity or caste), household (sex of household head, household economic status), geographic (rural or urban place of residence, subnational region) [22]. Using the WHO inequality work as a starting point, we conducted a systematic review to select the pertinent DHS variables for our inequity analysis. We conducted a systematic review to analyse qualitative and quantitative peer-reviewed literature of studies focused on inequities in childhood immunisation in low- and middle-income countries (LMICs) to identify the social determinants of childhood immunisation. Figure A2 (in appendix) illustrates the process flow diagram of identification, screening, eligibility, and inclusion of articles for the systematic review, using the PRISMA (Preferred Reporting Items for Systematic Reviews and Meta-Analyses) framework [23]. We conducted our search using MEDLINE and additional studies were identified through hand-searches of reference lists for articles written in the English language, published between 01/01/2010 to 04/10/2021, for which the full text was available, and contained the following terms in English or American spellings: (Vaccination coverage or Immunisation coverage) AND (Determinant, characteristic, predictor) AND (Equity, equality, disparity, inequality, inequity). We did not search prior to 2010 as papers and the subsequently identified social determinants may be less relevant to the current context. We identified 160 publications, screened the title and abstract, assessed full articles for eligibility, and included 49 publications in our systematic review (see Table 1). In addition to the characteristics analysed by the WHO inequality report on immunisation [22], we identified that antenatal care and birth setting had evidence of association with vaccination coverage.

**Table 1.**
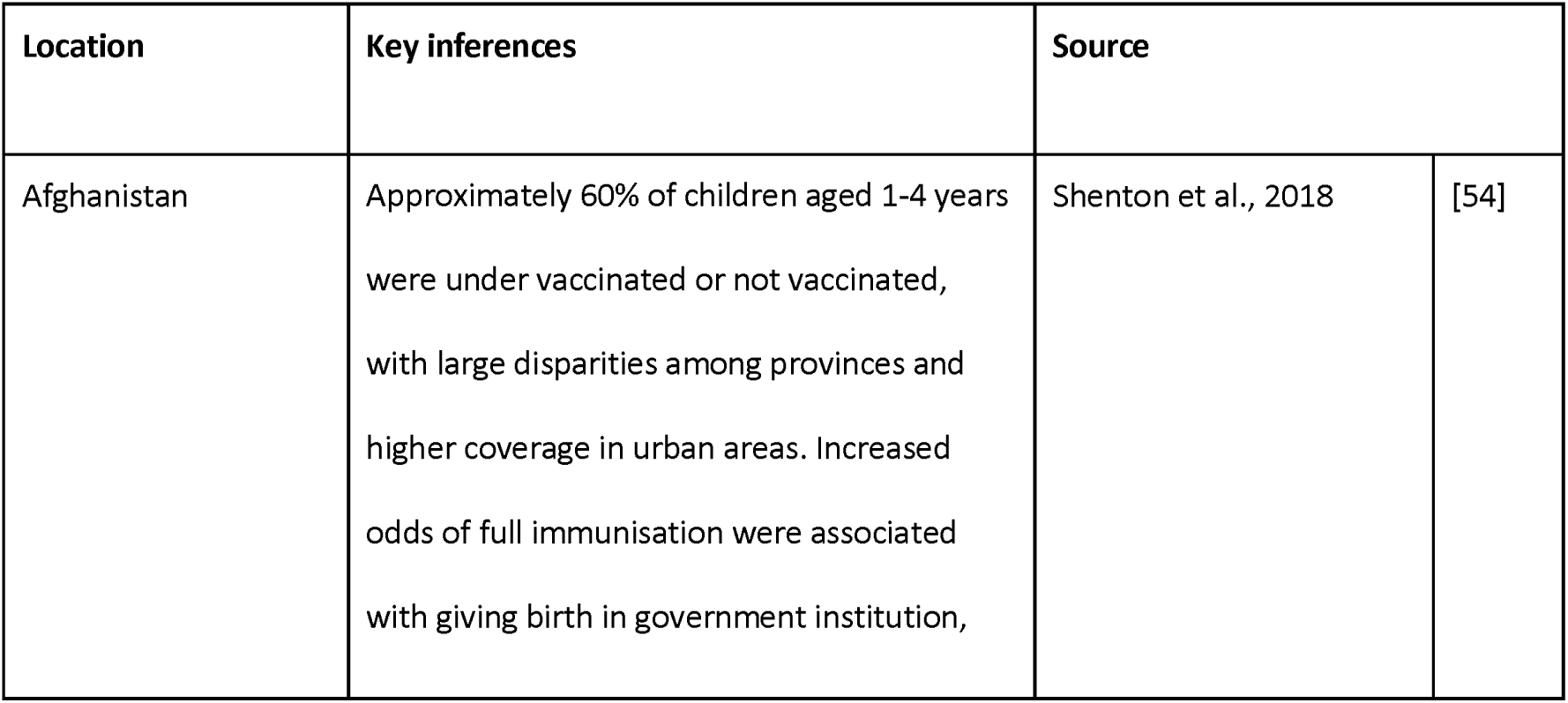
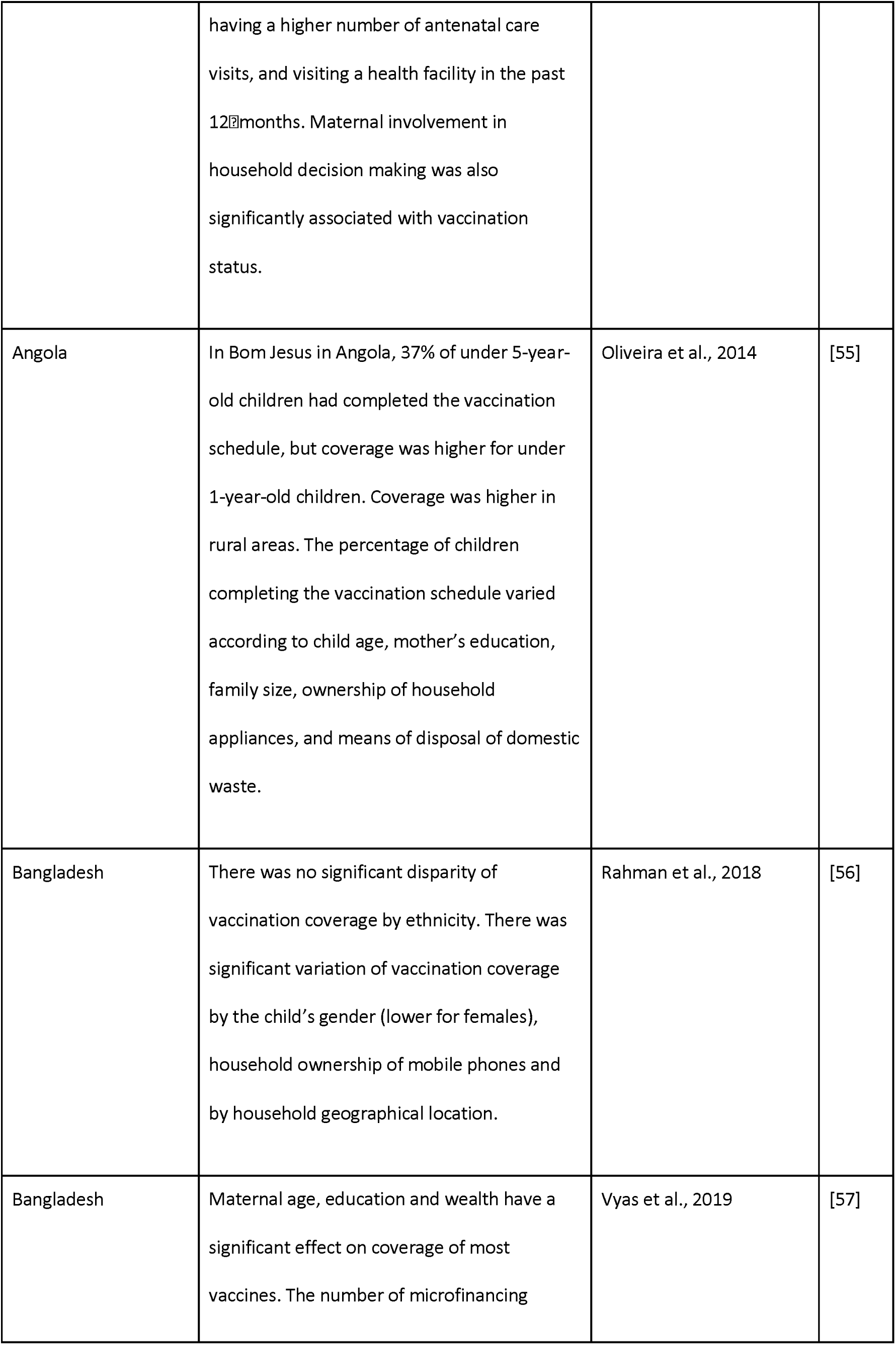
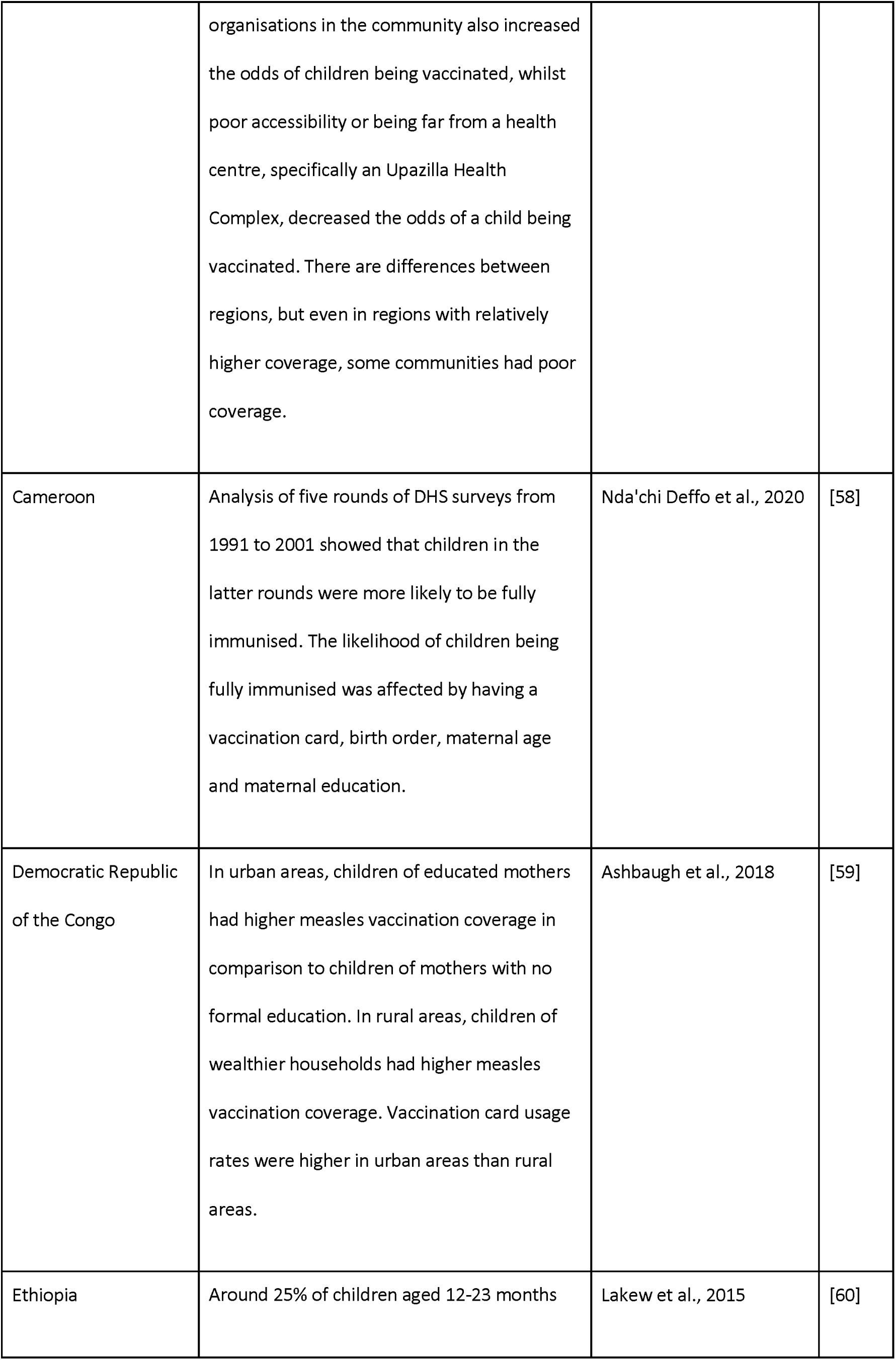
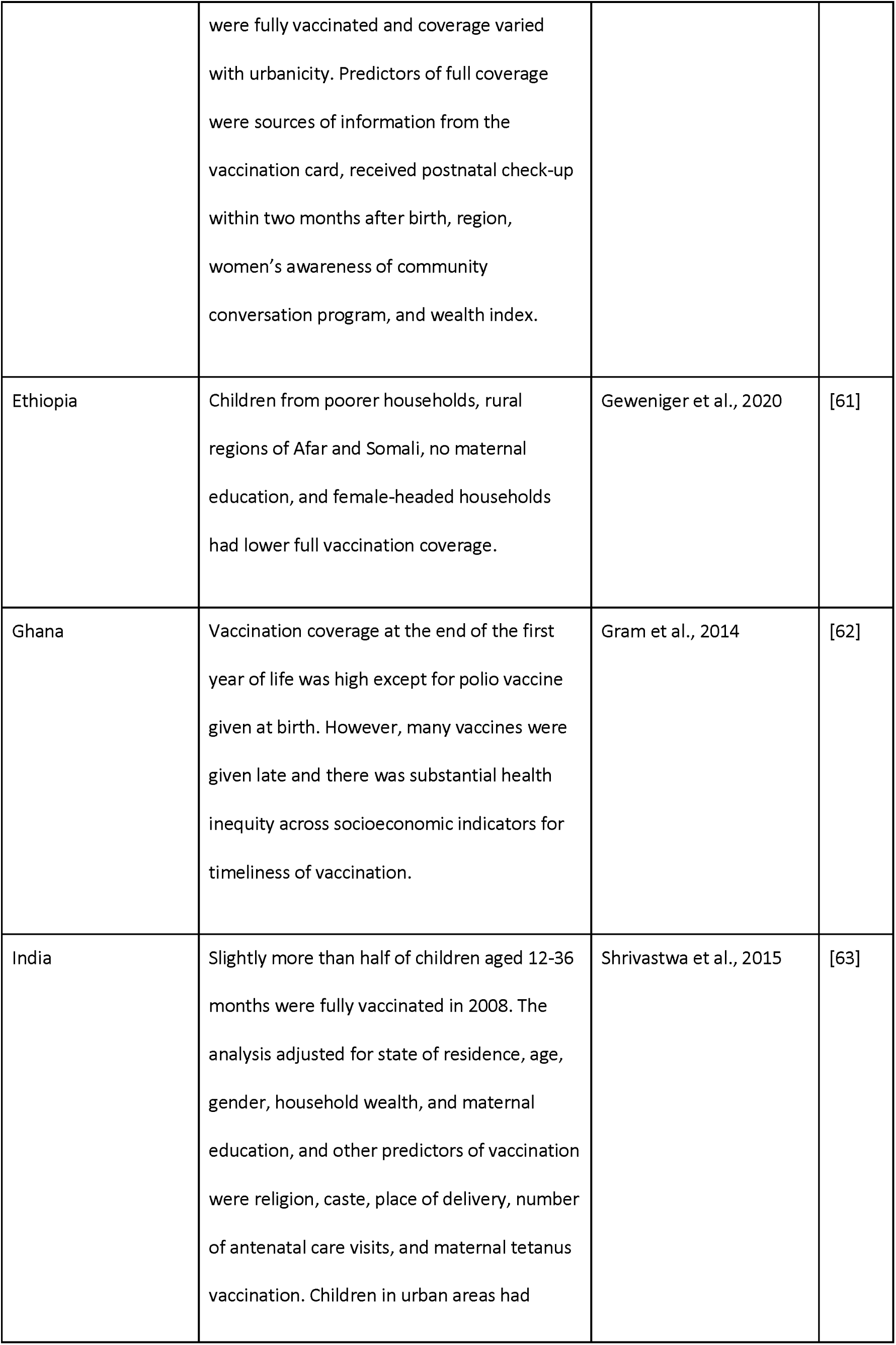
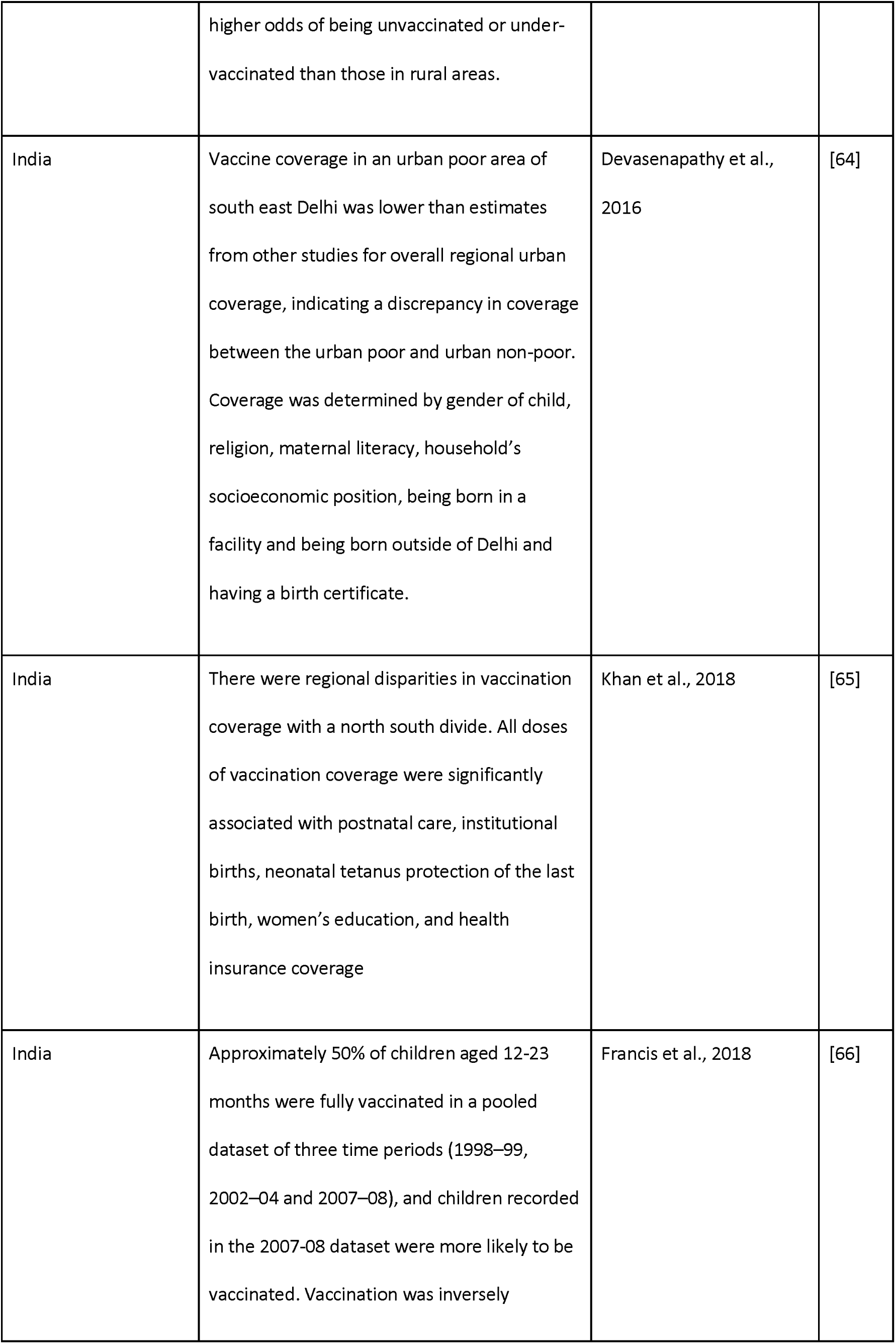
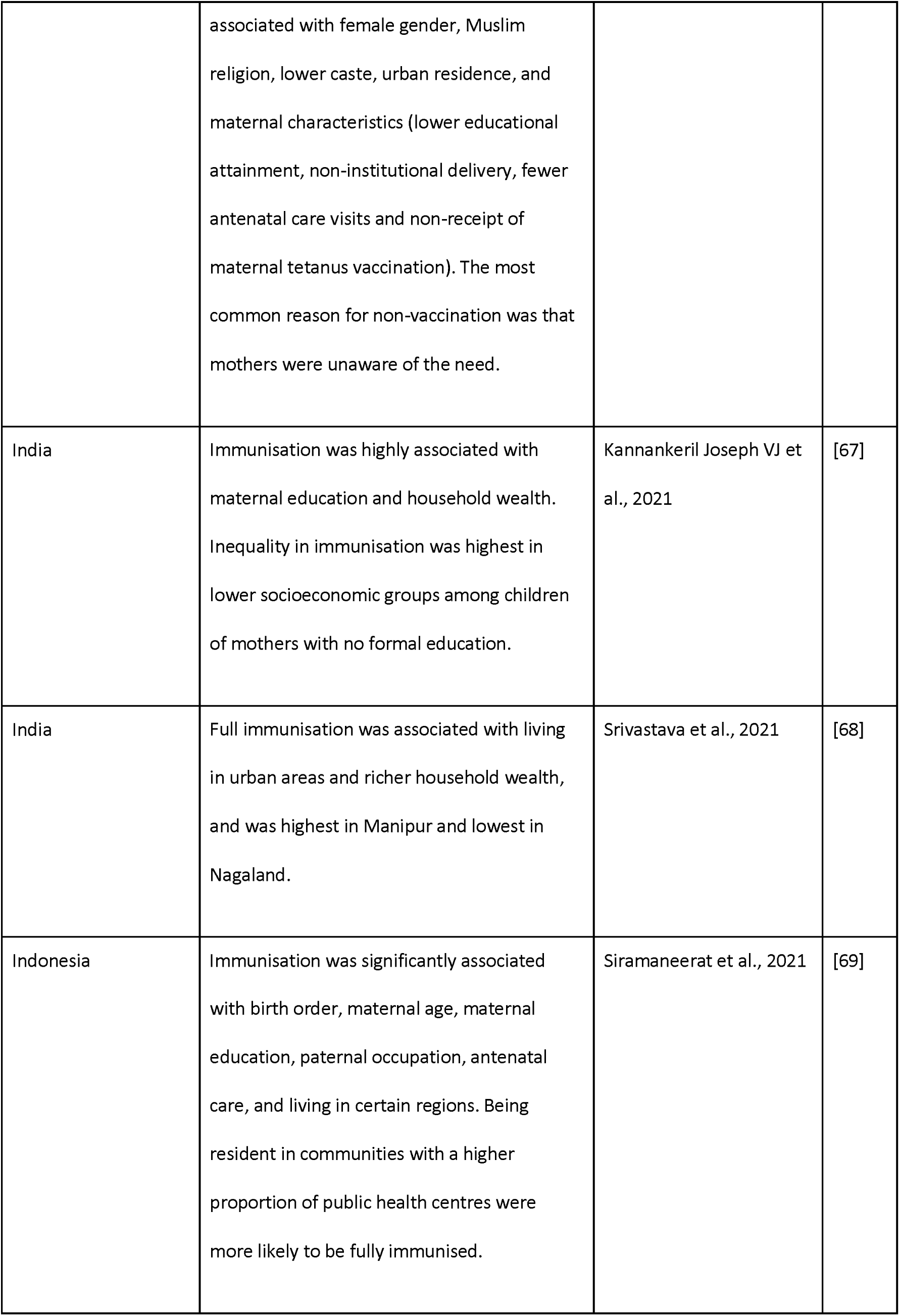
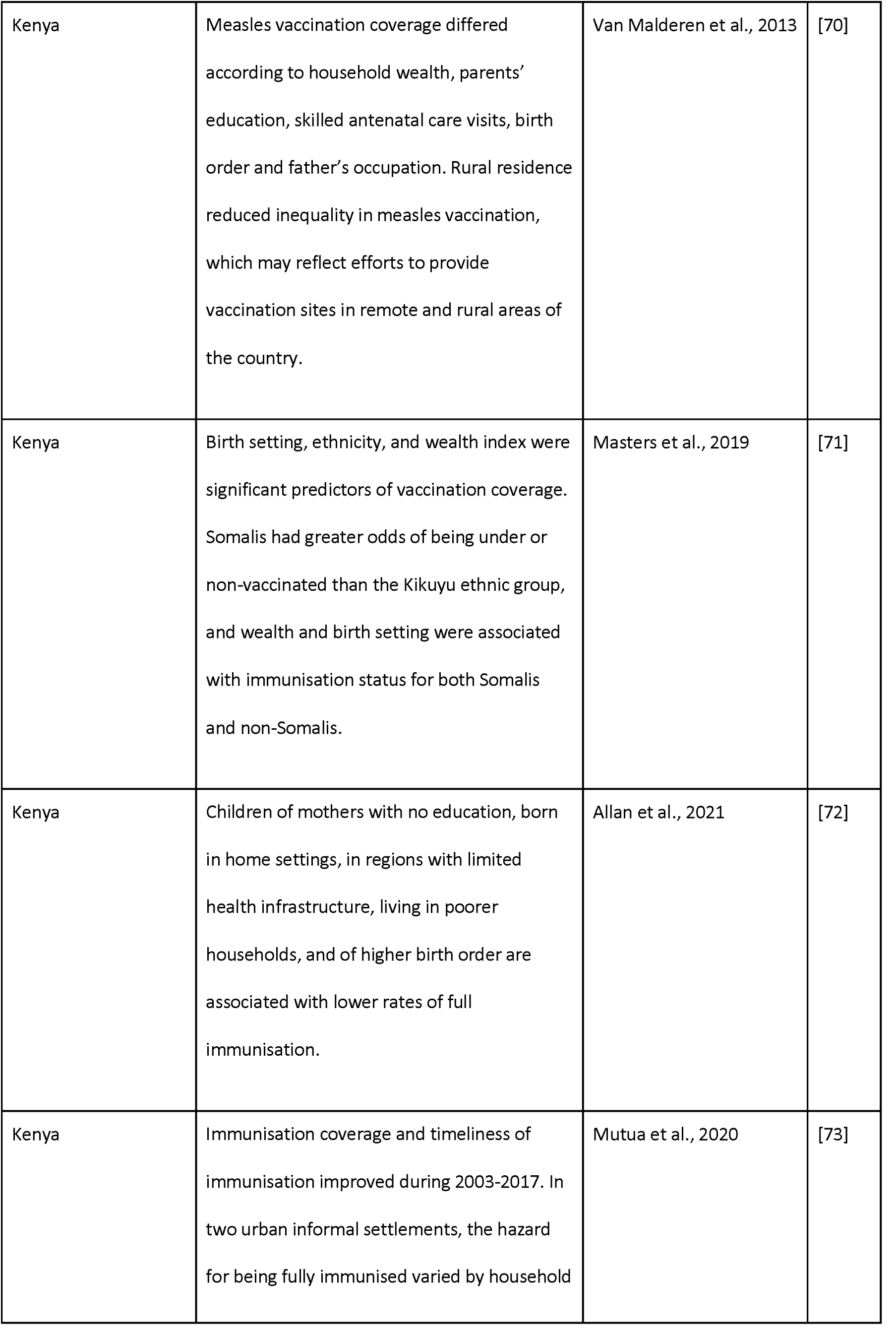
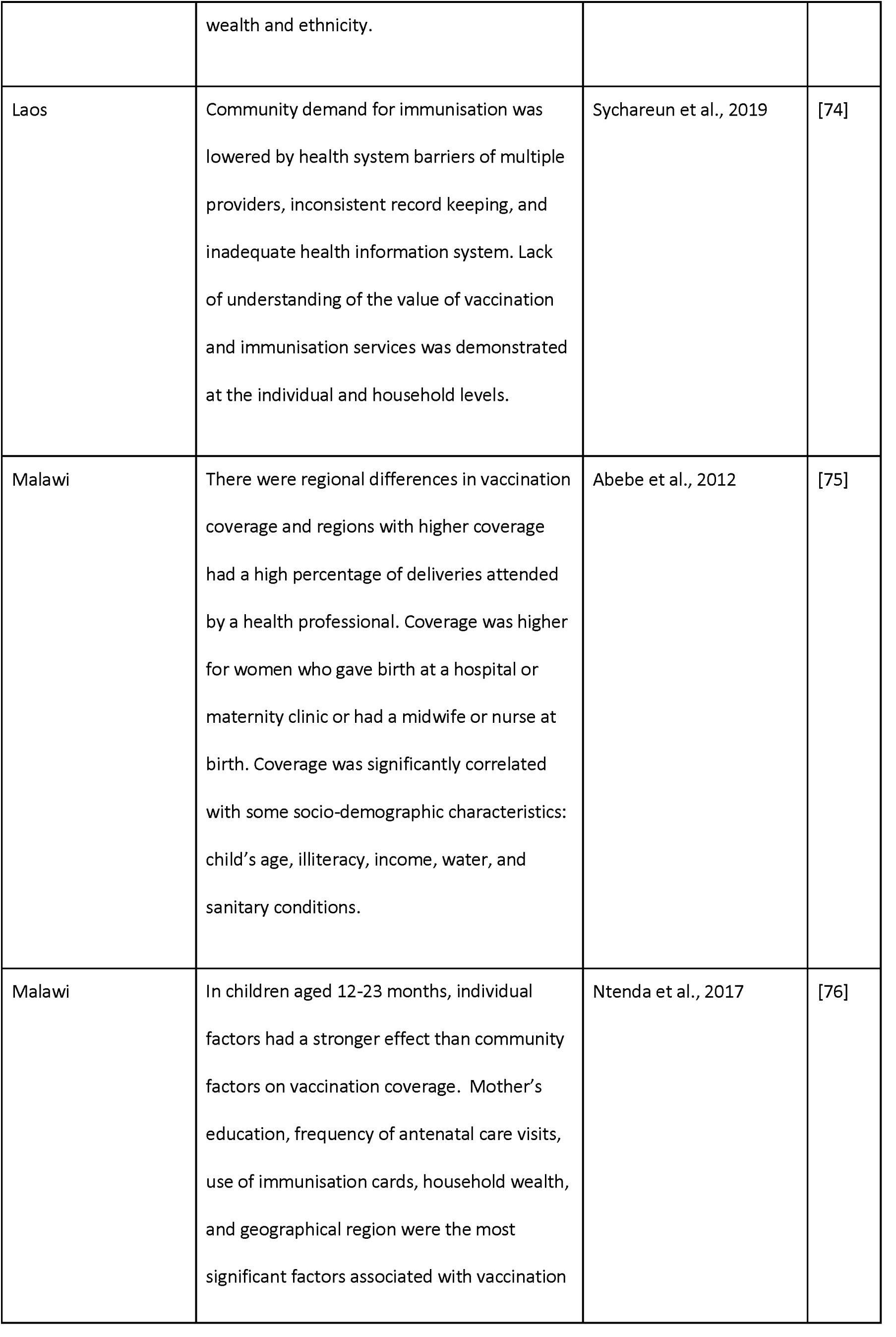
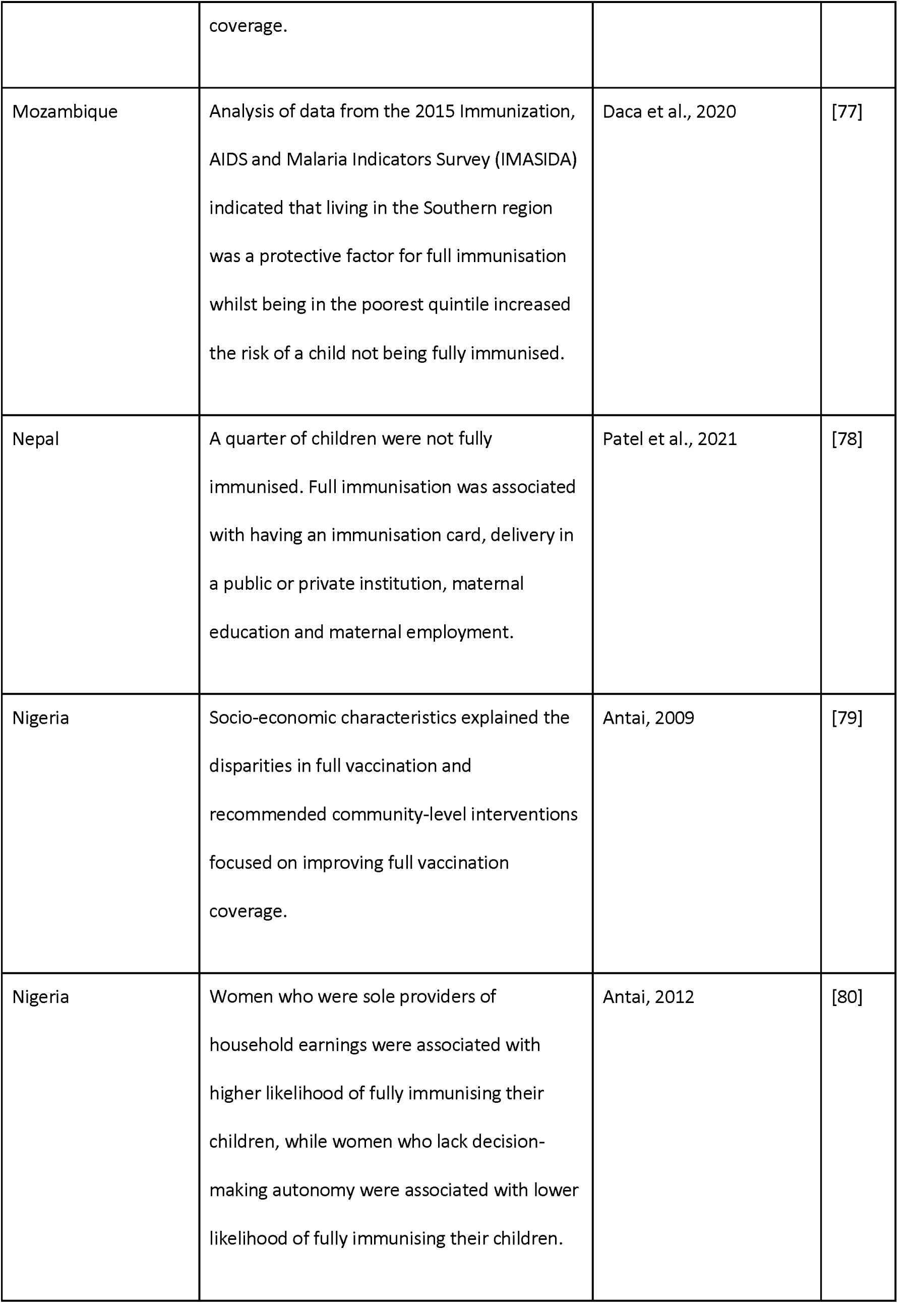
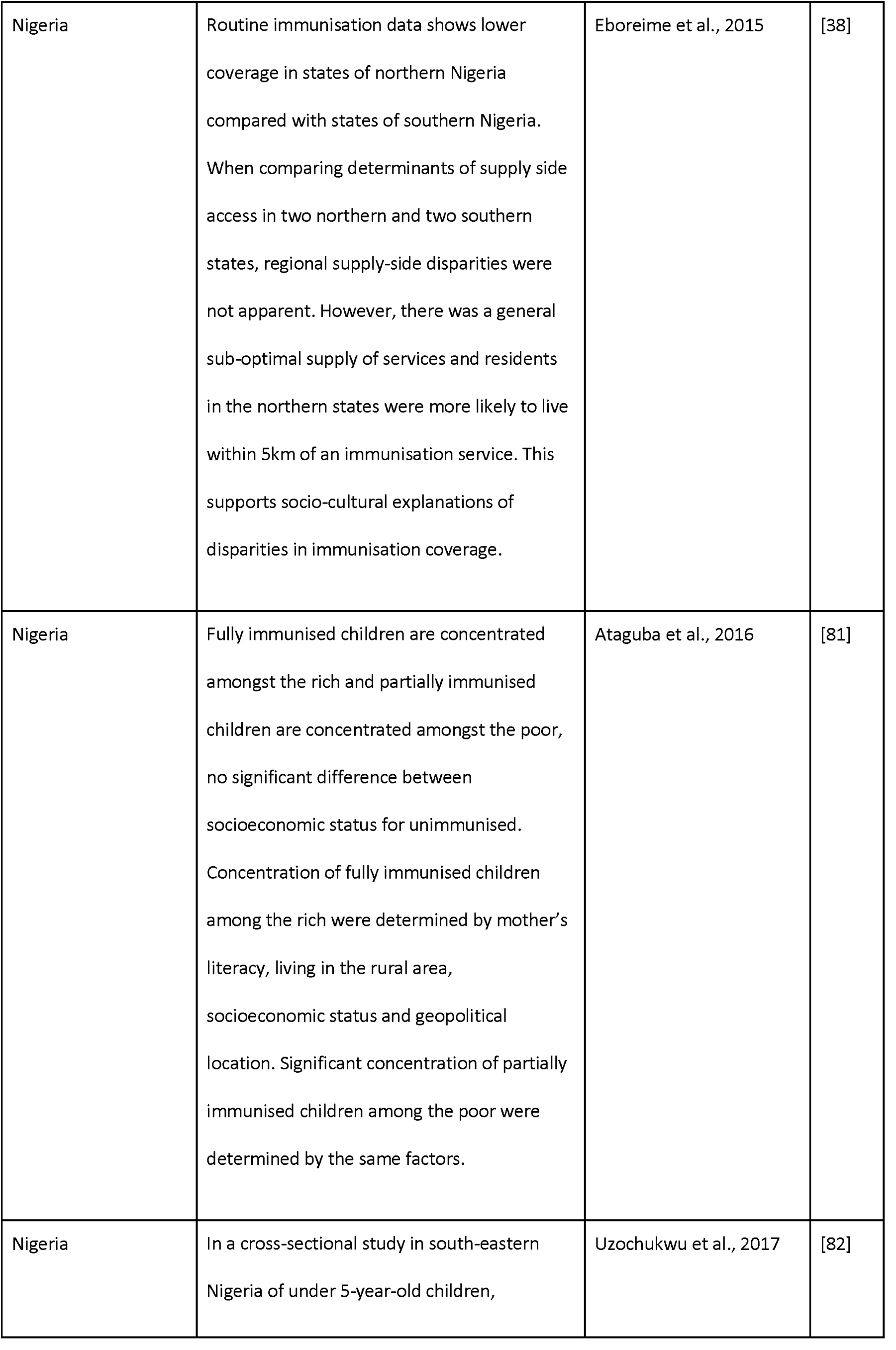
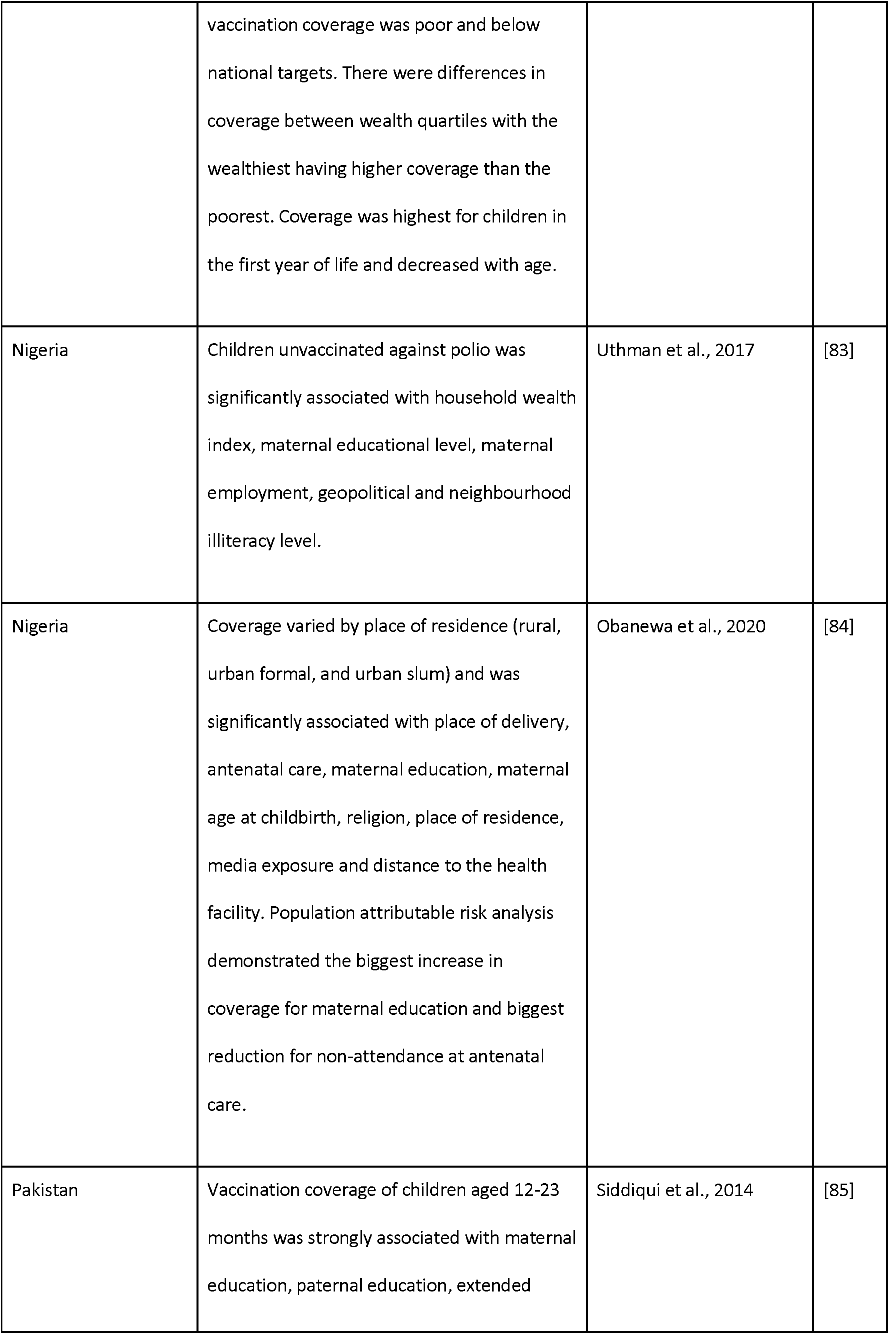
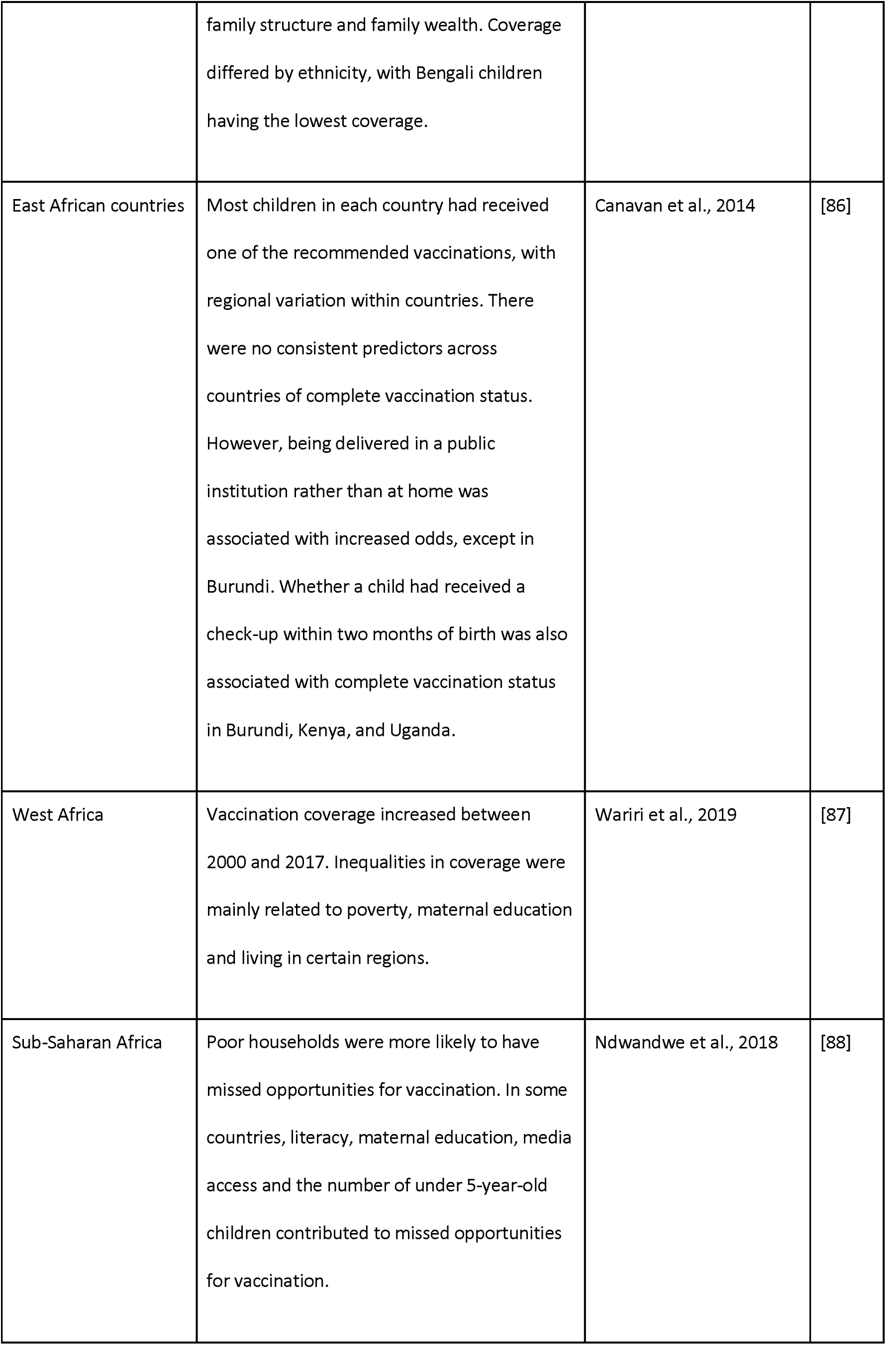
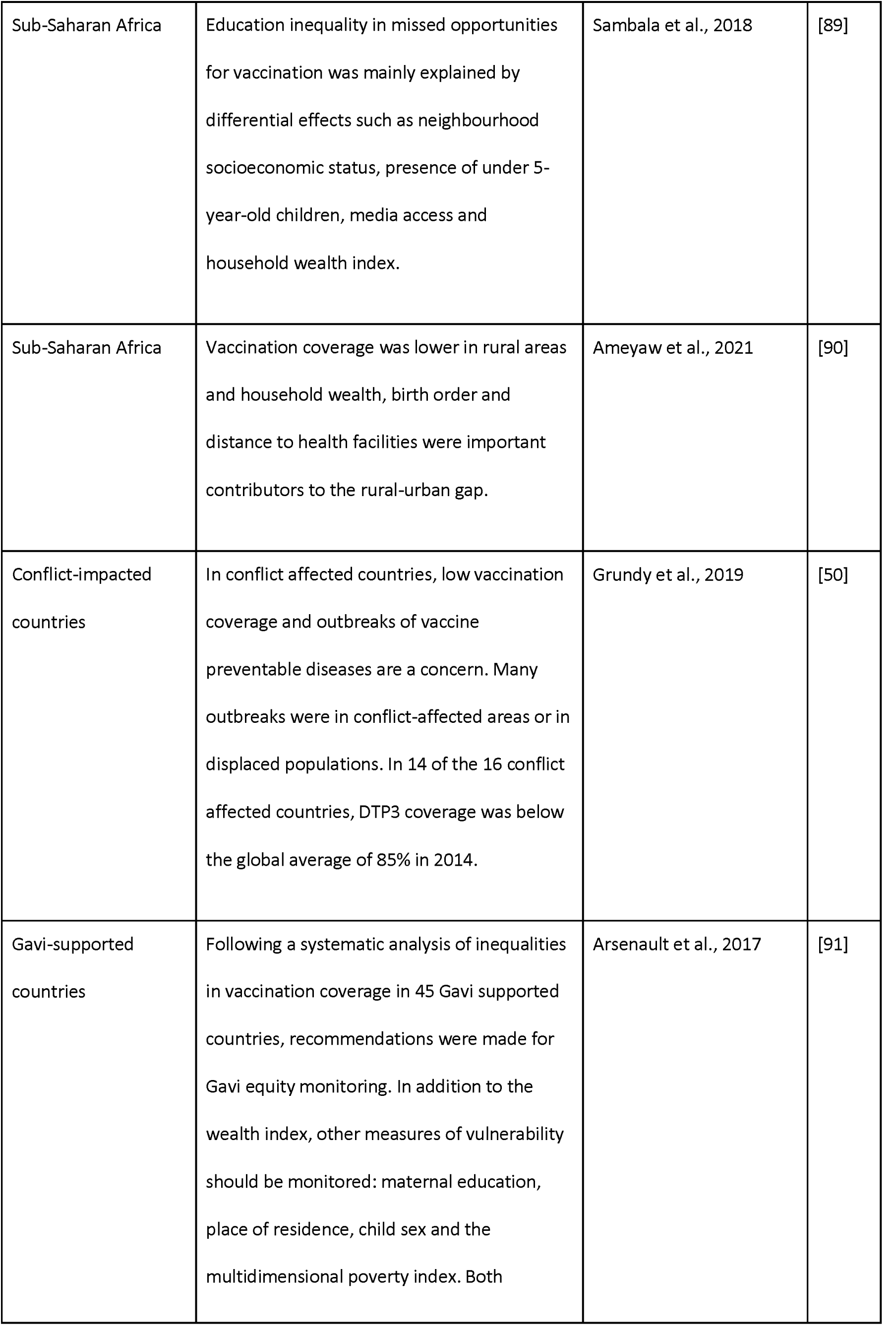
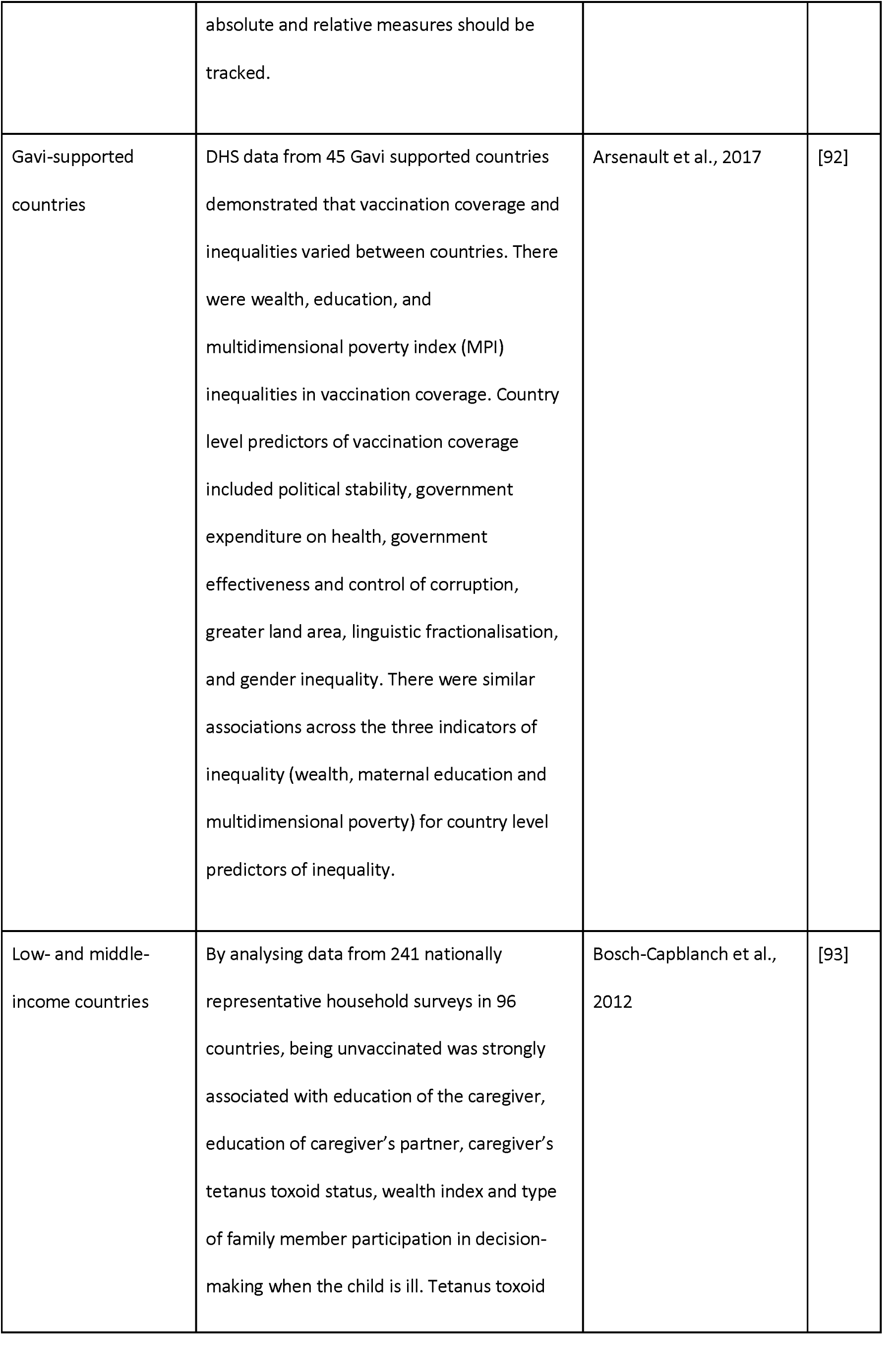
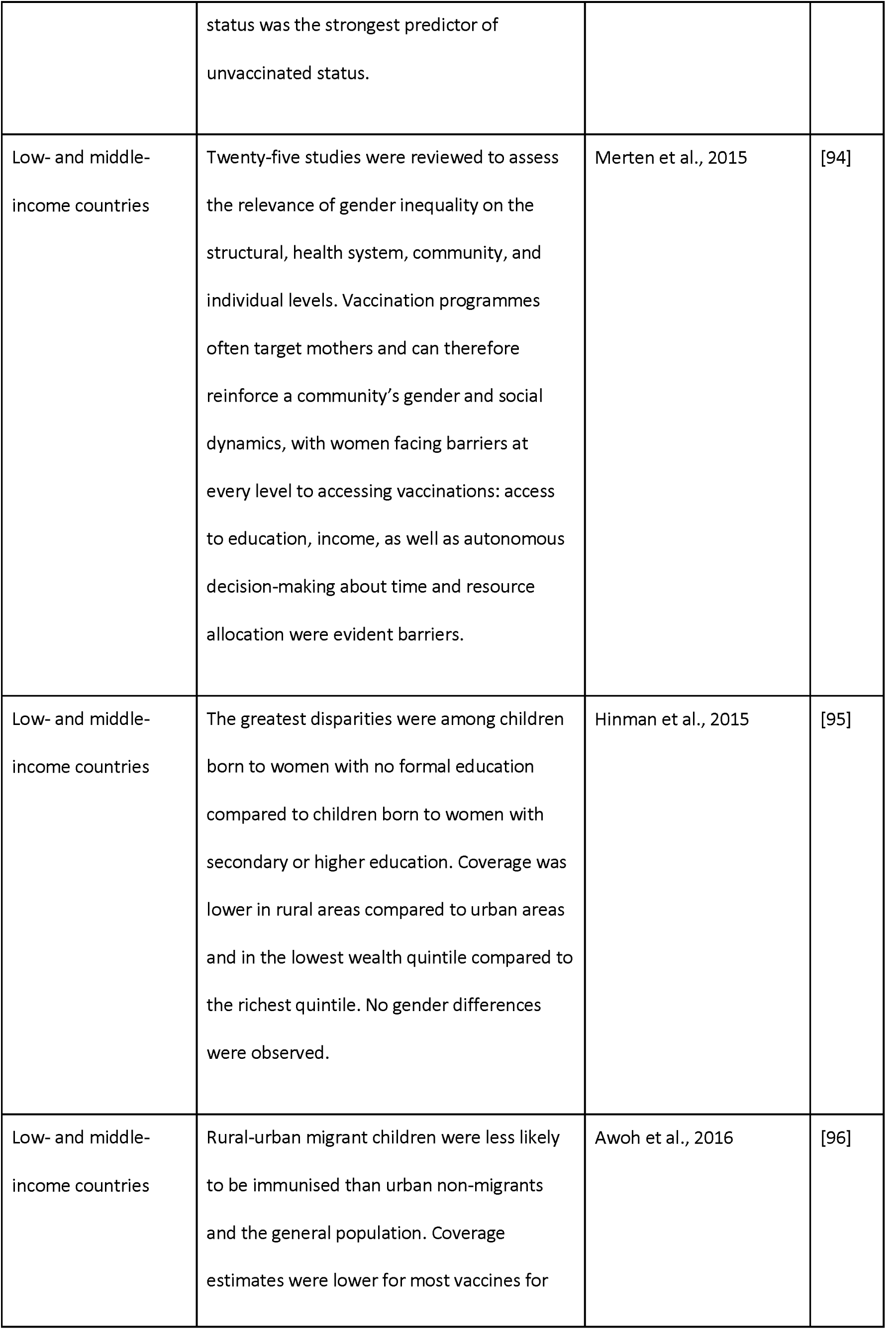
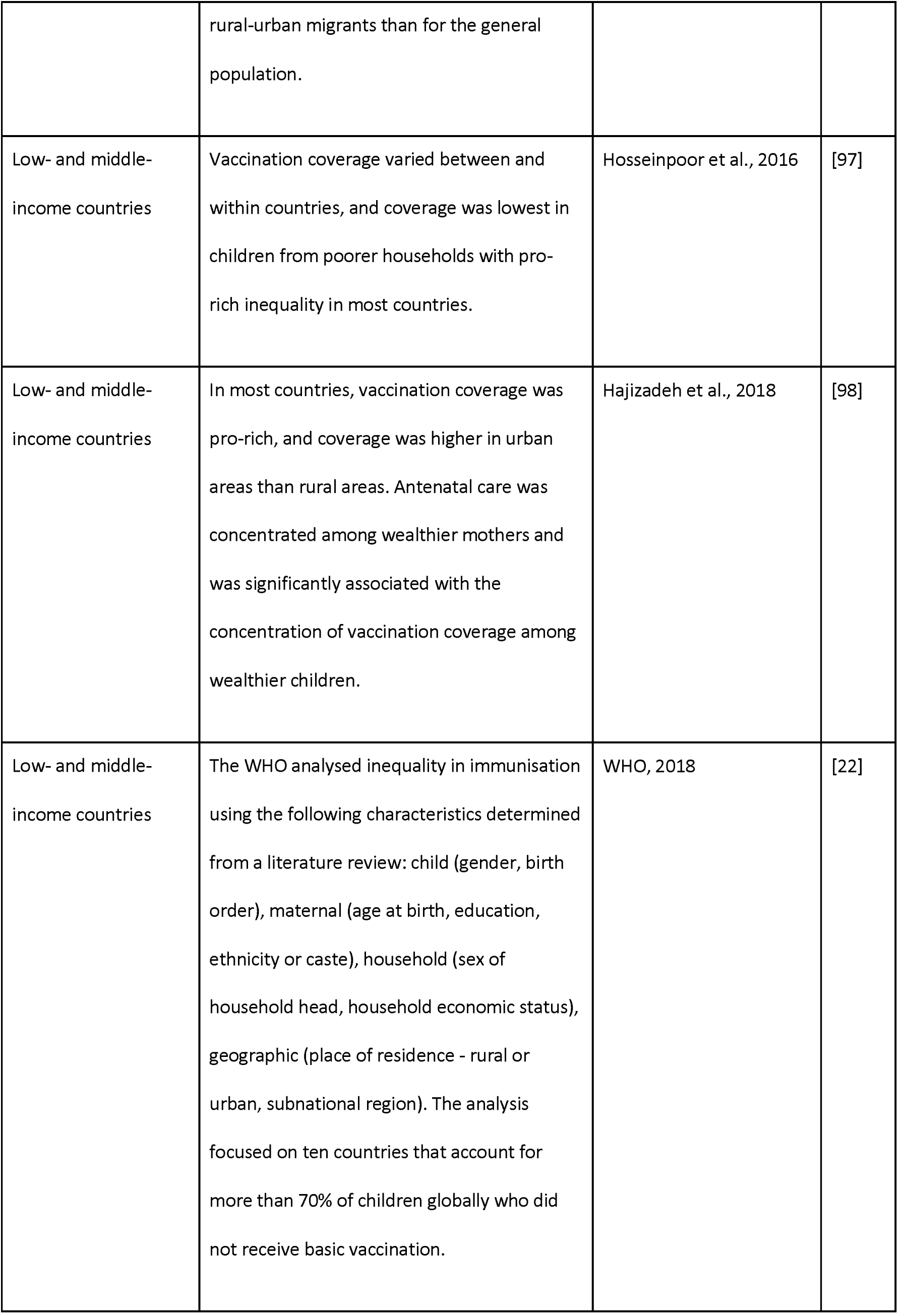
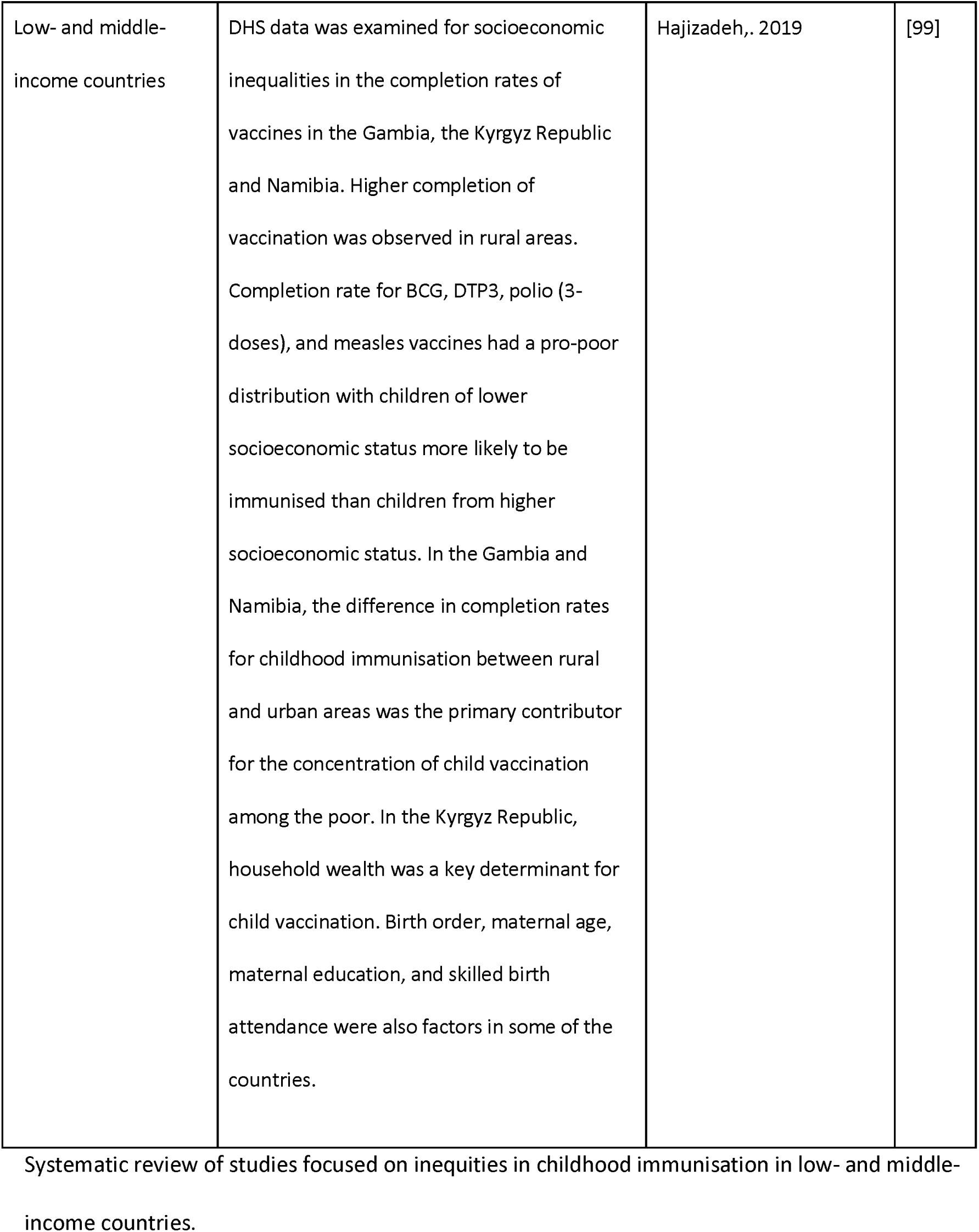
Systematic review of social determinants of childhood immunisation.

### Demographic and health survey

We analysed the 2018 Nigeria DHS which was conducted between August to December 2018 [24]. The DHS are nationally representative household surveys focusing on population, health, and nutrition in LMICs [25]. The DHS sample is a two-stage stratified cluster sample with sampling weights applied to ensure that results are representative. There are four questionnaires: household questionnaire, woman’s questionnaire, man’s questionnaire, and biomarker questionnaire. The country is divided into clusters with 30 households selected from each cluster. The woman’s questionnaire was asked to women aged 15-49 years and provides the data for our study. All women aged 15-49 years in the sampled households were included and the survey was successfully conducted in 1,389 clusters after 11 clusters were dropped following deteriorating security in those areas during data collection. In addition, in the state of Borno, 11 of the 27 Local Government Areas (LGAs) in the state were dropped due to insecurity. Clusters selected from these dropped LGAs were replaced with clusters from the remaining LGAs and so may not be representative of the entire state [24]. The study population for our analysis was women aged 15-49 years with a child aged 12 to 23 months old. Of the 41,821 women interviewed, 33,924 women had a child aged 59 months or younger and immunisation data was collected for 6,059 living children aged between 12-23 months. We applied sampling weights to the survey dataset to adjust for disproportionate sampling and non-response, thereby ensuring that the sample was representative of the population.

### Vaccination coverage

The information of whether a child has received a vaccination is gathered from the child’s vaccination card. If that is not available or if a vaccine has not been recorded, then the mother is asked which vaccines have been given to her child [24]. Our primary outcome and dependent variable of interest was basic vaccination coverage, that is the proportion of children receiving 1-dose BCG, 3-dose DTP-HepB-Hib, 3-dose polio, and 1-dose measles vaccines. DHS gives the vaccination status of “received” and “not received” for individual vaccines. Binary variables of “received all three doses” and “not received all three doses”were generated for the three dose vaccines. The basic vaccination variable was generated as a binary variable of “received all basic vaccinations” and “not received all basic vaccinations” by combining the variables for 1-dose BCG, 3-dose DTP-HepB-Hib, 3-dose polio, and 1-dose measles vaccines.

### Inequity analysis

We conducted simple logistic regression to estimate crude odds ratios and assess basic vaccination coverage disaggregated by socioeconomic, geographic, maternal, child, and healthcare characteristics as measured by the DHS and identified through the systematic review and the WHO inequality report on immunisation [22]. Due to the survey design, a p-value for all categories within a characteristic was determined using the Adjusted Wald’s test through the “test” function in Stata [26]. These p-values were used to assess the association between basic vaccination coverage and the characteristics [27] and if a p-value was <0.05 then the characteristic was included in the model. We checked for collinearity between variables and avoided multicollinearity in the development of a parsimonious model [27] using the “regress” in Stata and calculation of the variance inflation factor. However, no variables required removal from the model.

Interaction was tested using the “contrast” function in Stata. This gives F statistics which are adjusted for the survey design, with the p-value demonstrating the statistical significance of the interaction. As in the WHO report [22, 27], interactions were tested between mother’s education and household wealth, mother’s age and household wealth, mother’s age and education, and place of residence and household wealth. We conducted multivariable logistic regression to estimate adjusted odds ratios (AORs) for socioeconomic, geographic, maternal, child, and healthcare characteristics associated with basic vaccination coverage (see Table 3 for the characteristics included in the multivariable logistic regression and Tables A4 in the appendix for the strata specific AORs between place of residence and household wealth).

**Table 2.**
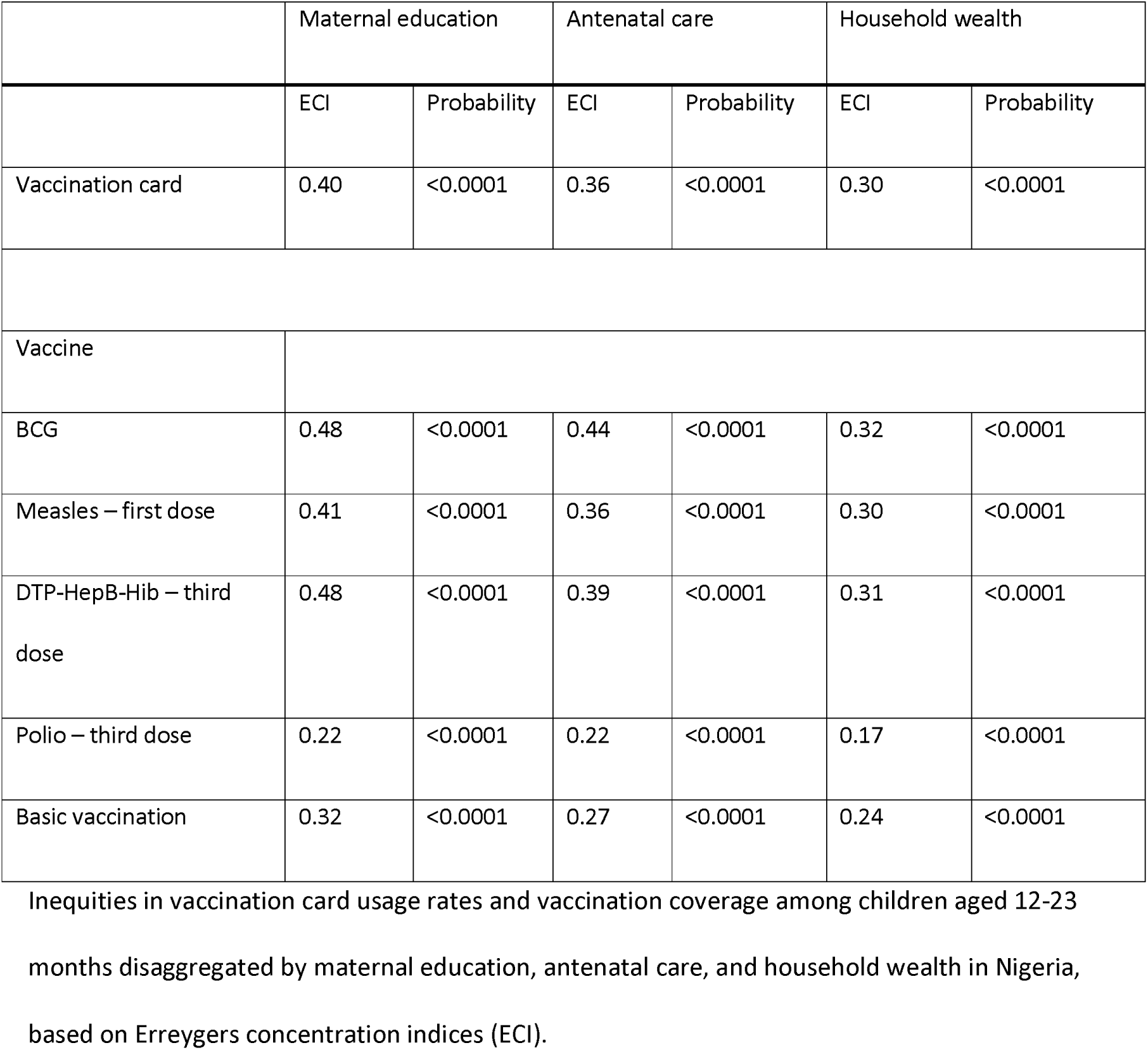
Inequities in vaccination card usage rates and vaccination coverage in Nigeria.

|  | Maternal education |  | Antenatal care |  | Household wealth |  |
| --- | --- | --- | --- | --- | --- | --- |
|  | ECI | Probability | ECI | Probability | ECI | Probability |
| Vaccination card | 0.40 | <0.0001 | 0.36 | <0.0001 | 0.30 | <0.0001 |
| Vaccine |  |  |  |  |  |  |
| BCG | 0.48 | <0.0001 | 0.44 | <0.0001 | 0.32 | <0.0001 |
| Measles – first dose | 0.41 | <0.0001 | 0.36 | <0.0001 | 0.30 | <0.0001 |
| DTP-HepB-Hib – third dose | 0.48 | <0.0001 | 0.39 | <0.0001 | 0.31 | <0.0001 |
| Polio – third dose | 0.22 | <0.0001 | 0.22 | <0.0001 | 0.17 | <0.0001 |
| Basic vaccination | 0.32 | <0.0001 | 0.27 | <0.0001 | 0.24 | <0.0001 |

Inequities in vaccination card usage rates and vaccination coverage among children aged 12-23
months disaggregated by maternal education, antenatal care, and household wealth in Nigeria,
based on Erreygers concentration indices (ECI).

**Table 3.**
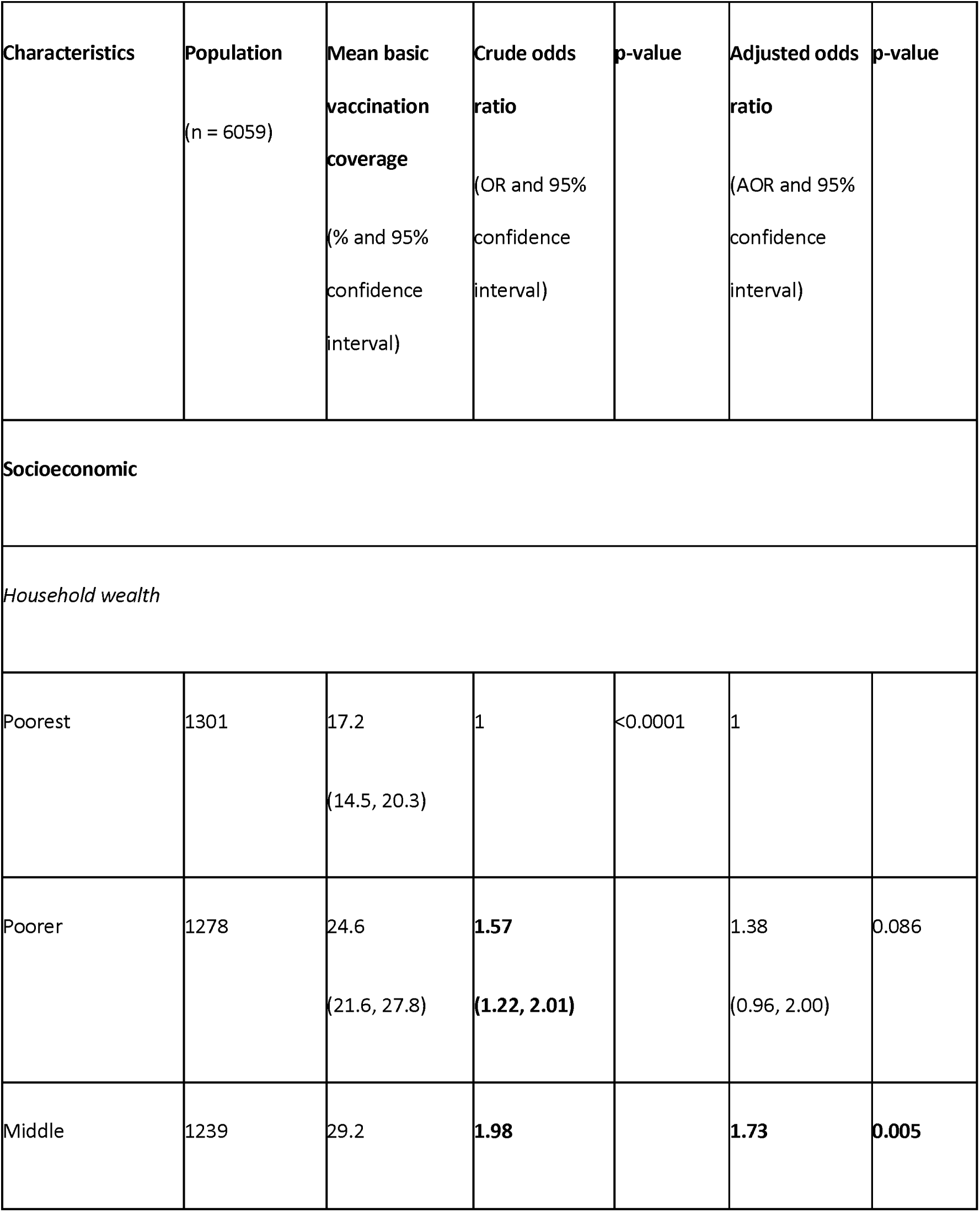
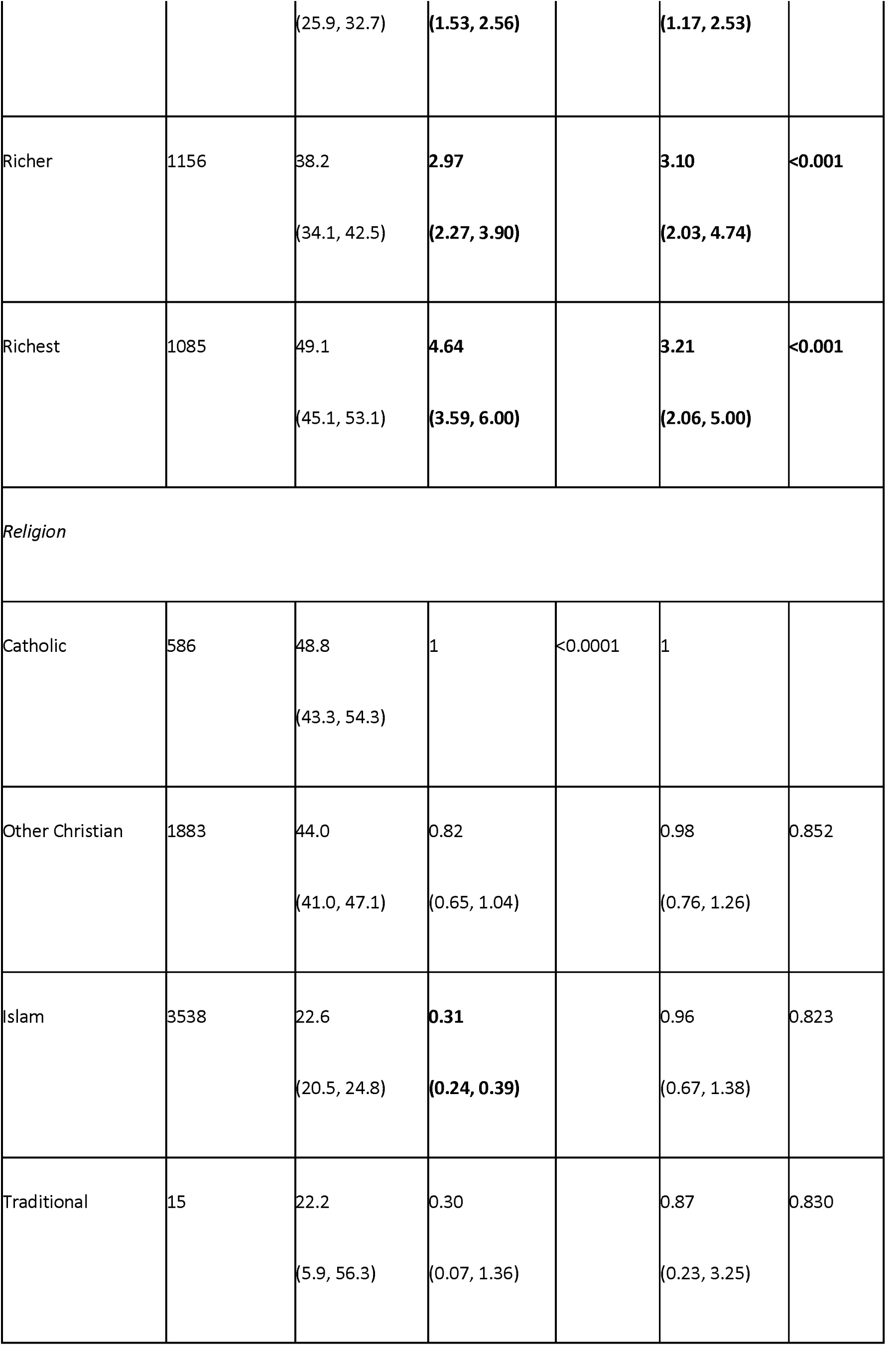
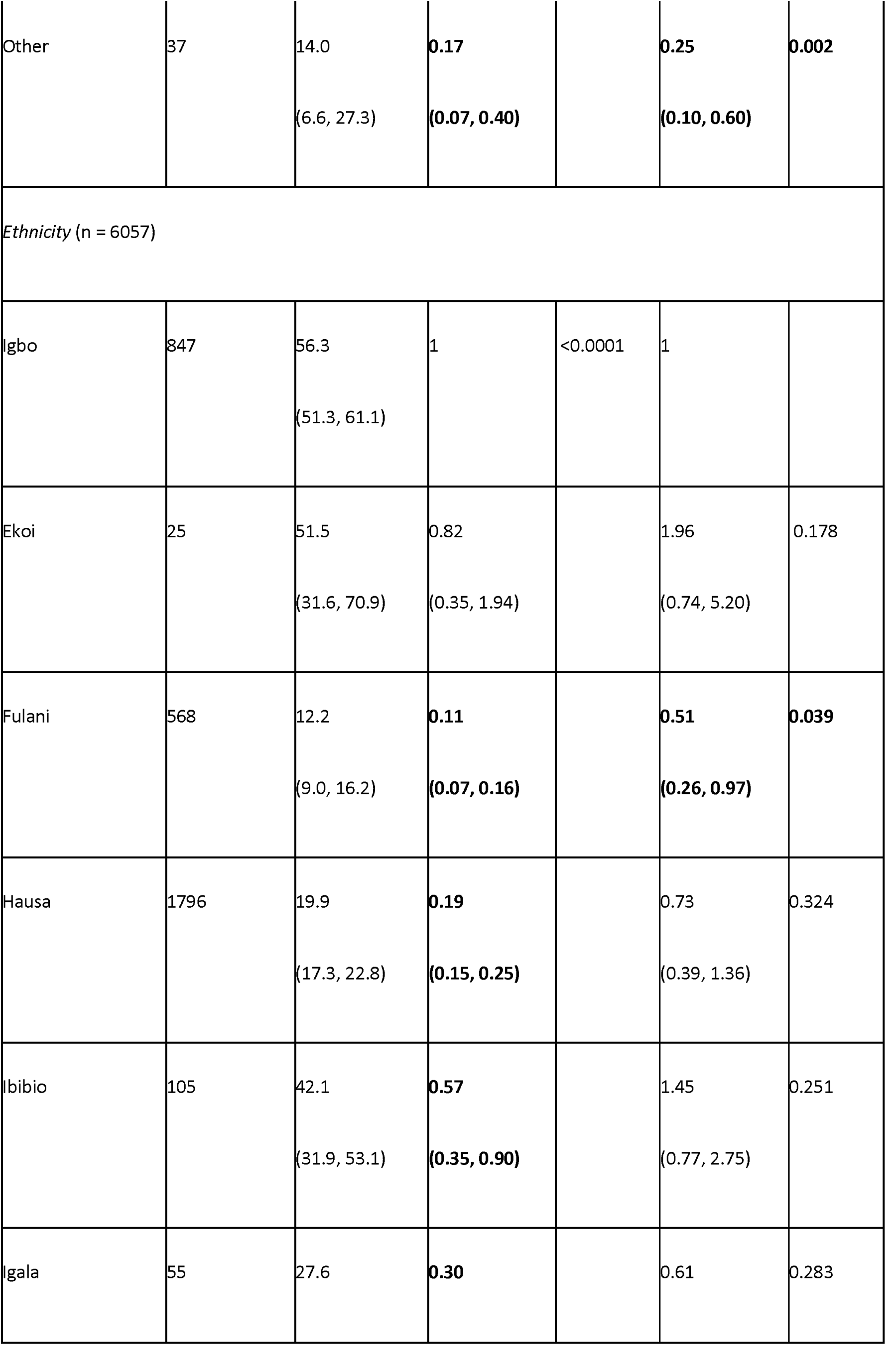
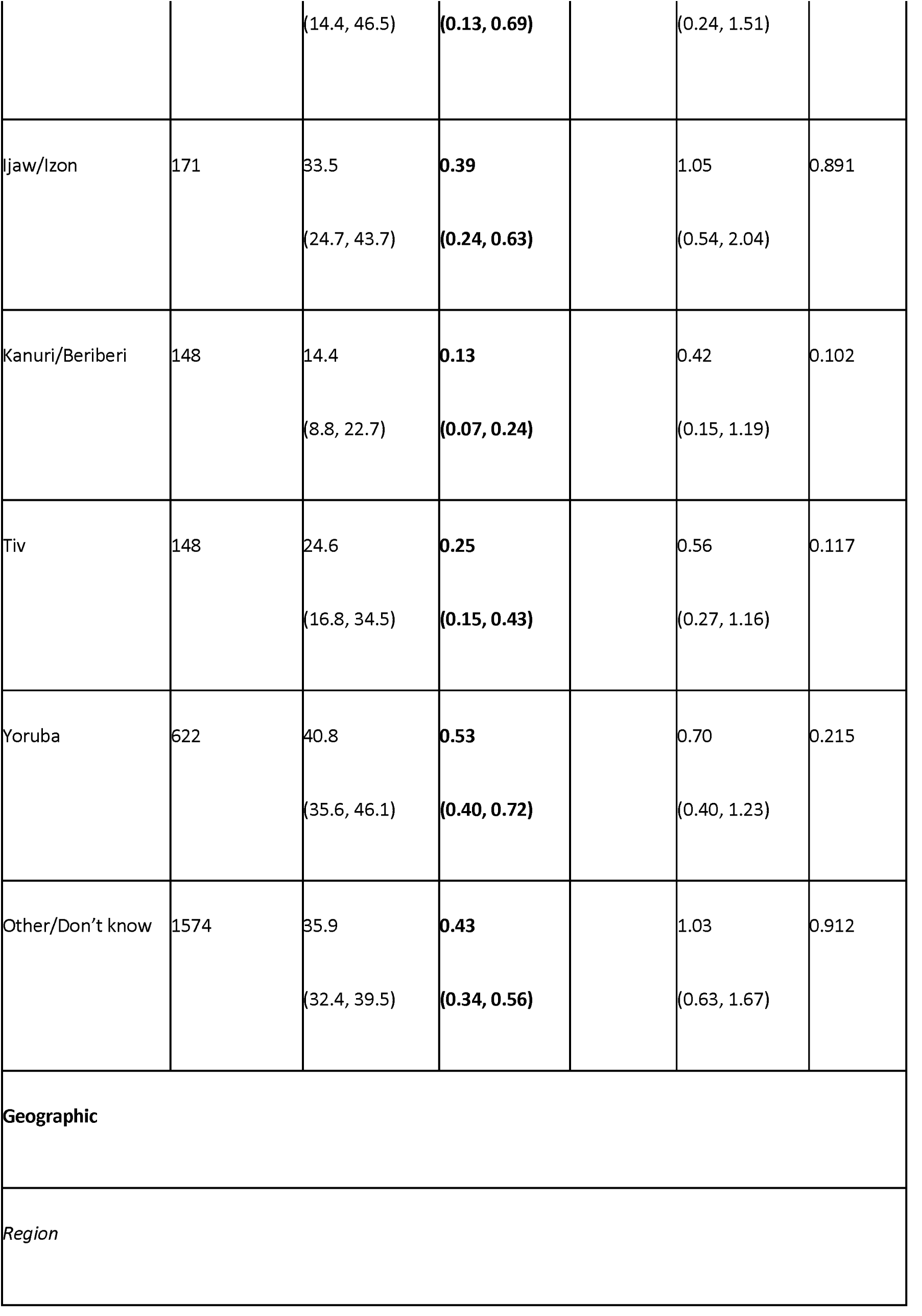
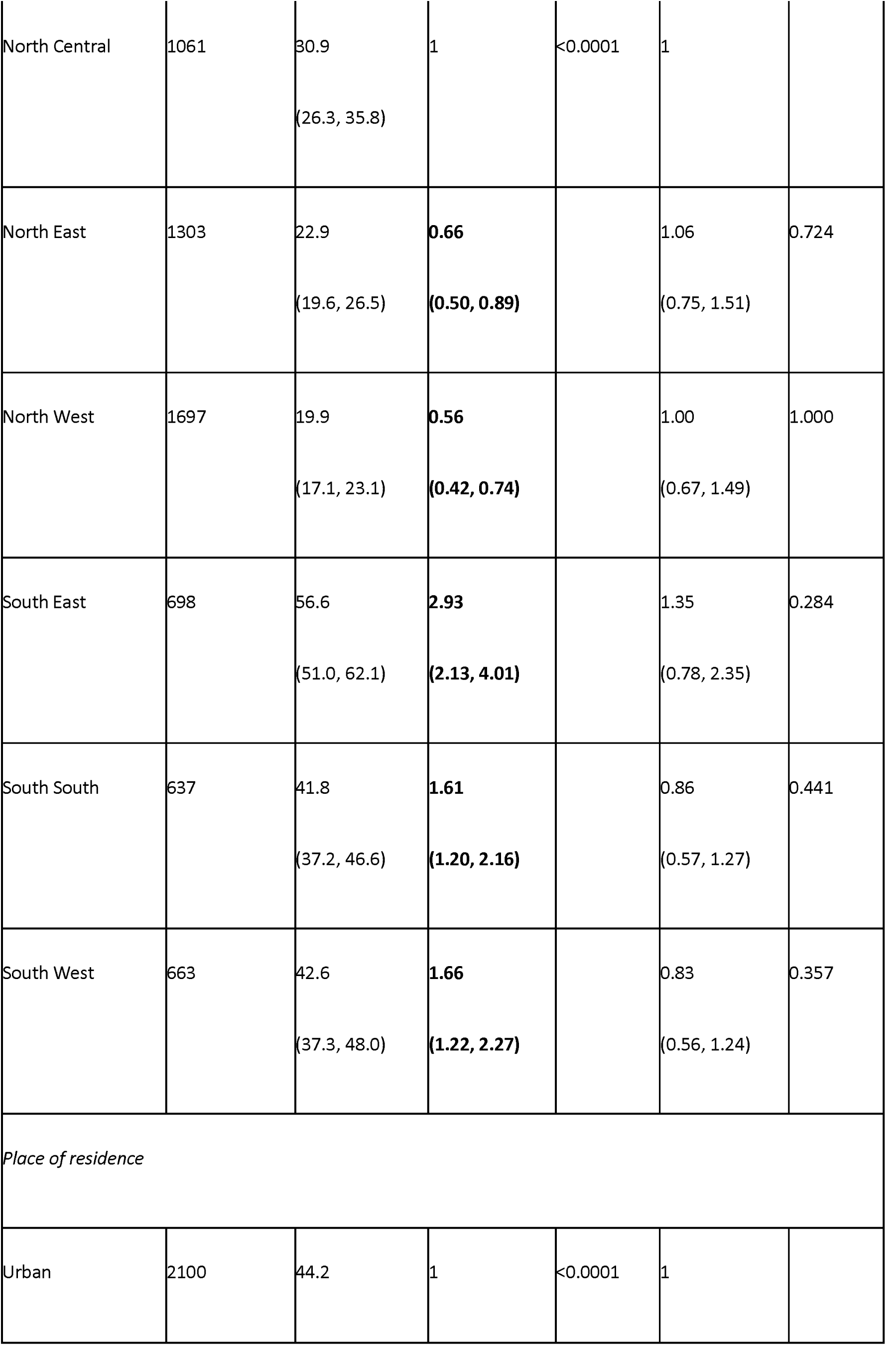
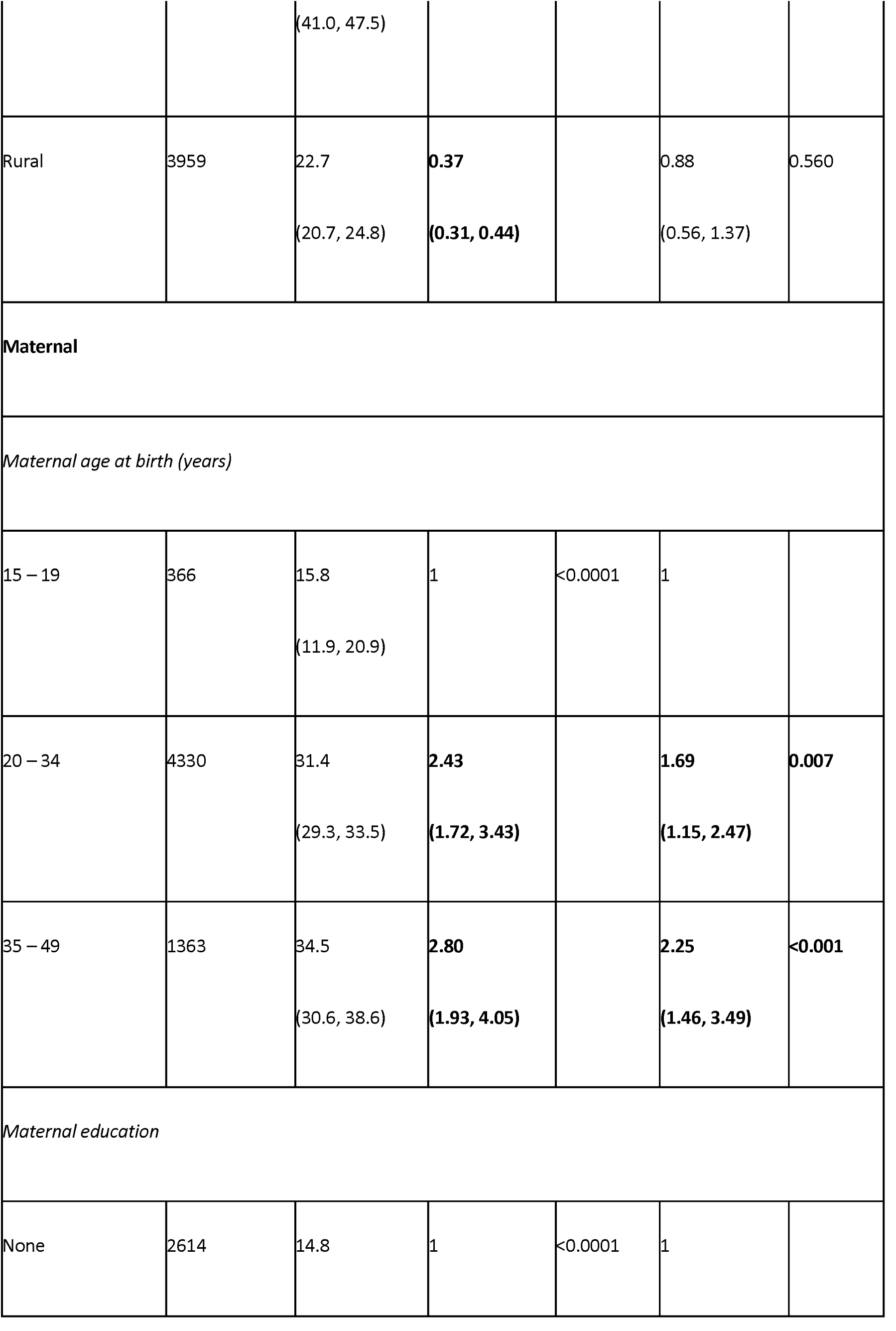
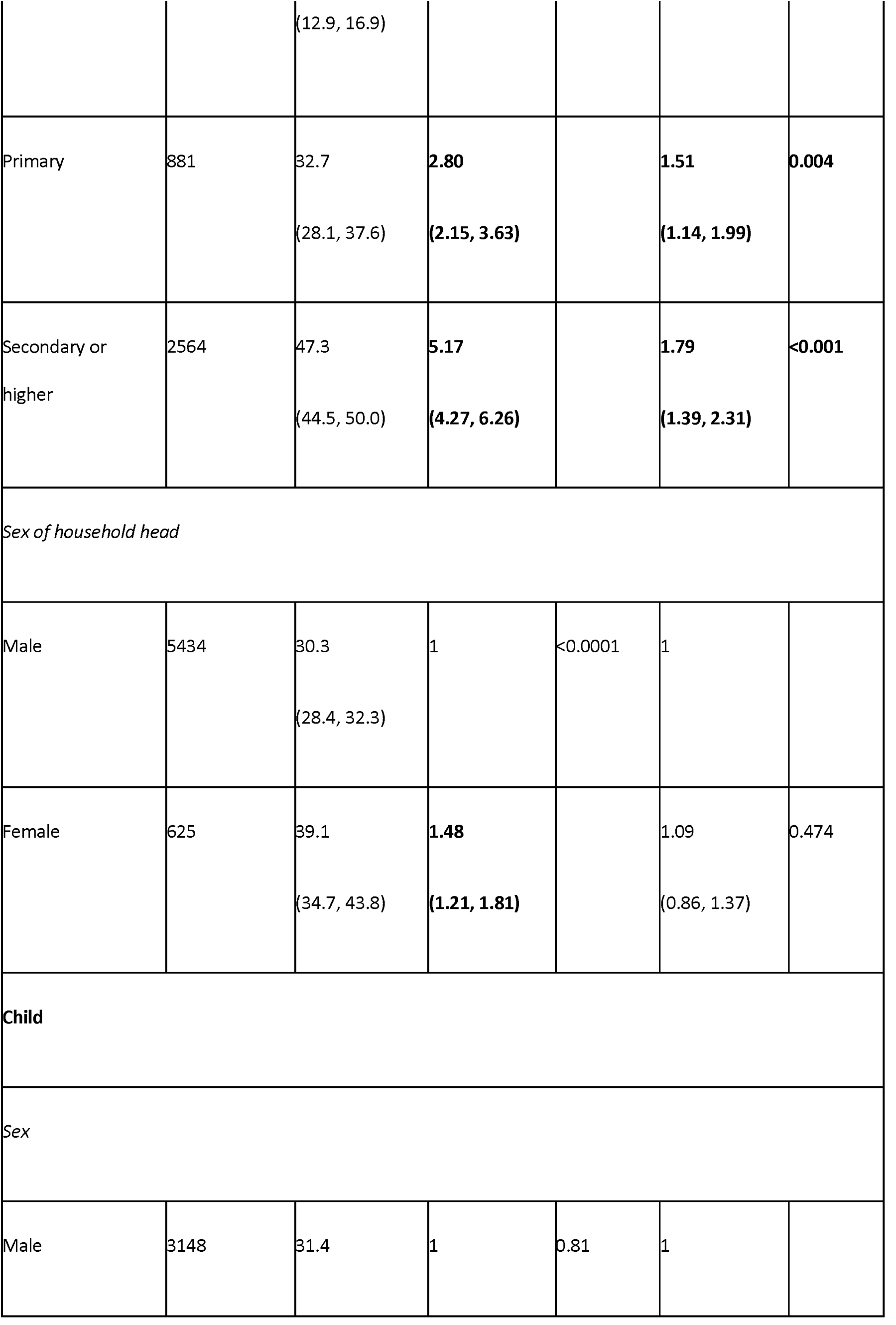
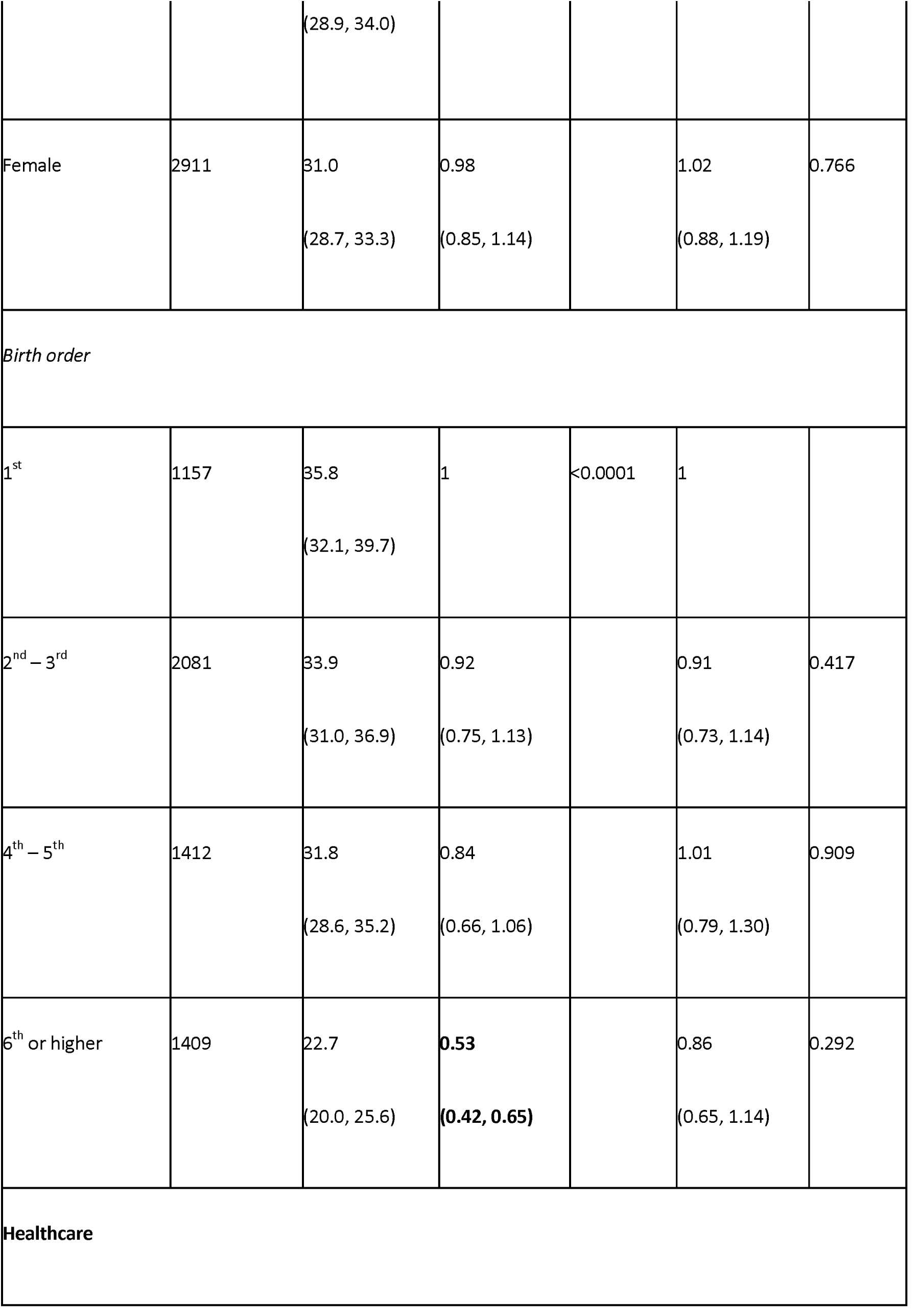
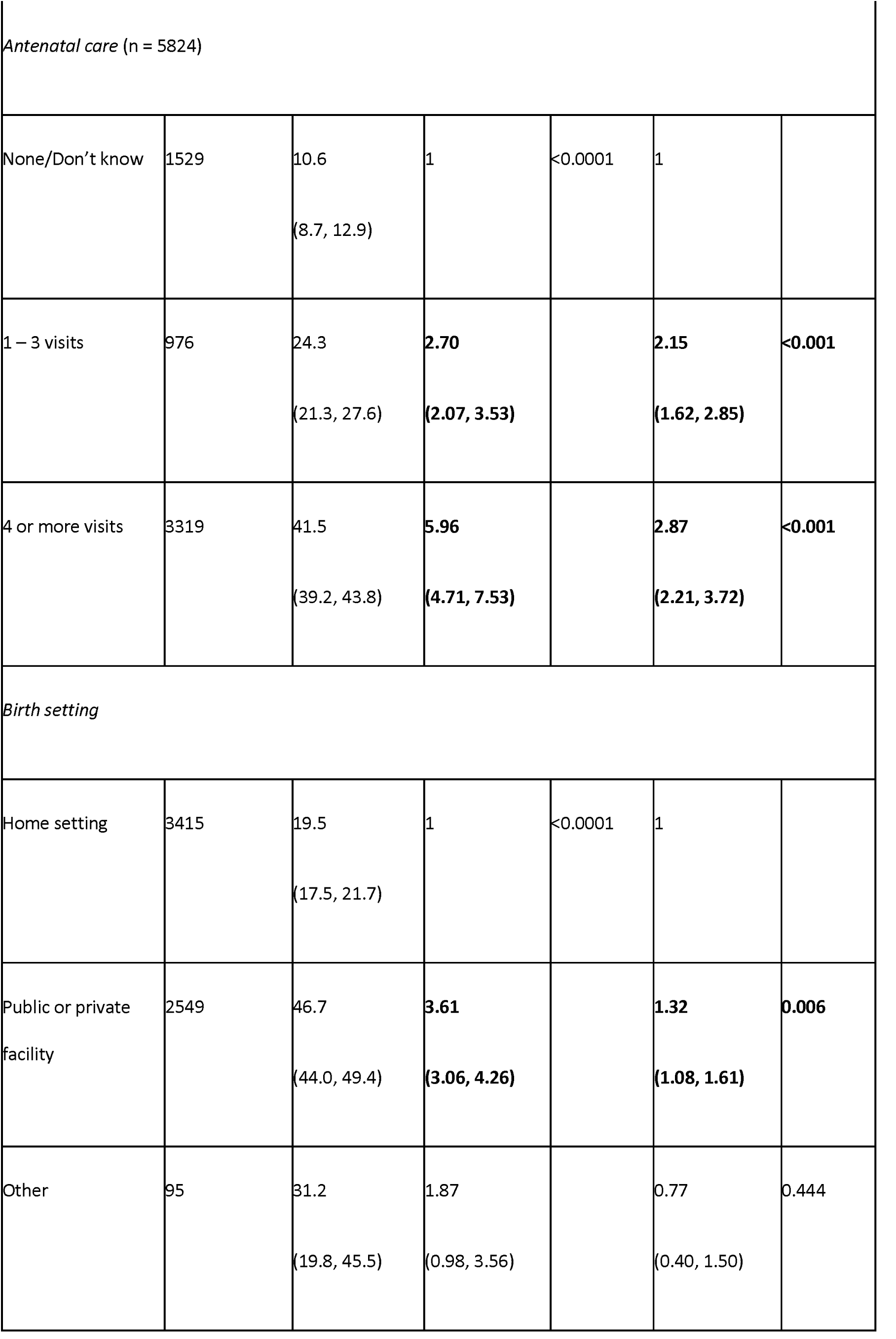
Inequities in basic vaccination coverage in Nigeria associated with socioeconomic, geographic, maternal, child, and healthcare characteristics.

We analysed inequity further by estimating the Erreygers concentration indices for maternal education, antenatal care, and household wealth to assess if basic vaccination coverage and vaccination card usage had progressive, regressive, or equal distribution based on each of these characteristics. The concentration index value shows how much of a health measure is concentrated in an advantaged or disadvantaged group. Values range from +1 to -1, with a value of zero meaning there is no inequity and positive values indicating that a health measure is concentrated in the more advantaged groups [28].

### Reproducibility of analysis

We conducted the survey analysis using the Stata statistical software [26], and visualisations were generated using the R statistical software [29]. The analysis code is publicly accessible on GitHub at https://github.com/svwilliams122/vaccine-equity-Nigeria and the 2018 Nigeria DHS data set is accessible upon registration on the DHS website at https://dhsprogram.com/methodology/survey/survey-display-528.cfm.

## Results

Through the systematic review for characteristics selection (see Table 1), we streamlined the social determinants of childhood immunisation to: household wealth, religion, and ethnicity for socioeconomic characteristics; region and place of residence for geographic characteristics; maternal age at birth, maternal education, and maternal household head status for maternal characteristics; sex of child and birth order for child characteristics; and antenatal care and birth setting for healthcare characteristics.

### Vaccination coverage

Among the 6,059 children aged 12-23 months in the 2018 Nigeria DHS following the application of sample weights, 2,100 (35%) of them lived in urban areas. The coverage for the individual vaccines of BCG, DTP-HepB-Hib, polio, and measles was higher in urban areas compared to rural areas (Figure 2 and Table A3). At the national level, the mean basic vaccination coverage was 31% (95% CI: 29-33) with coverage of single vaccinations ranging from 48% (46-50%) for the third dose of the polio vaccine to 73% (71-75%) for the first dose of the polio vaccine.

**Figure 2.**
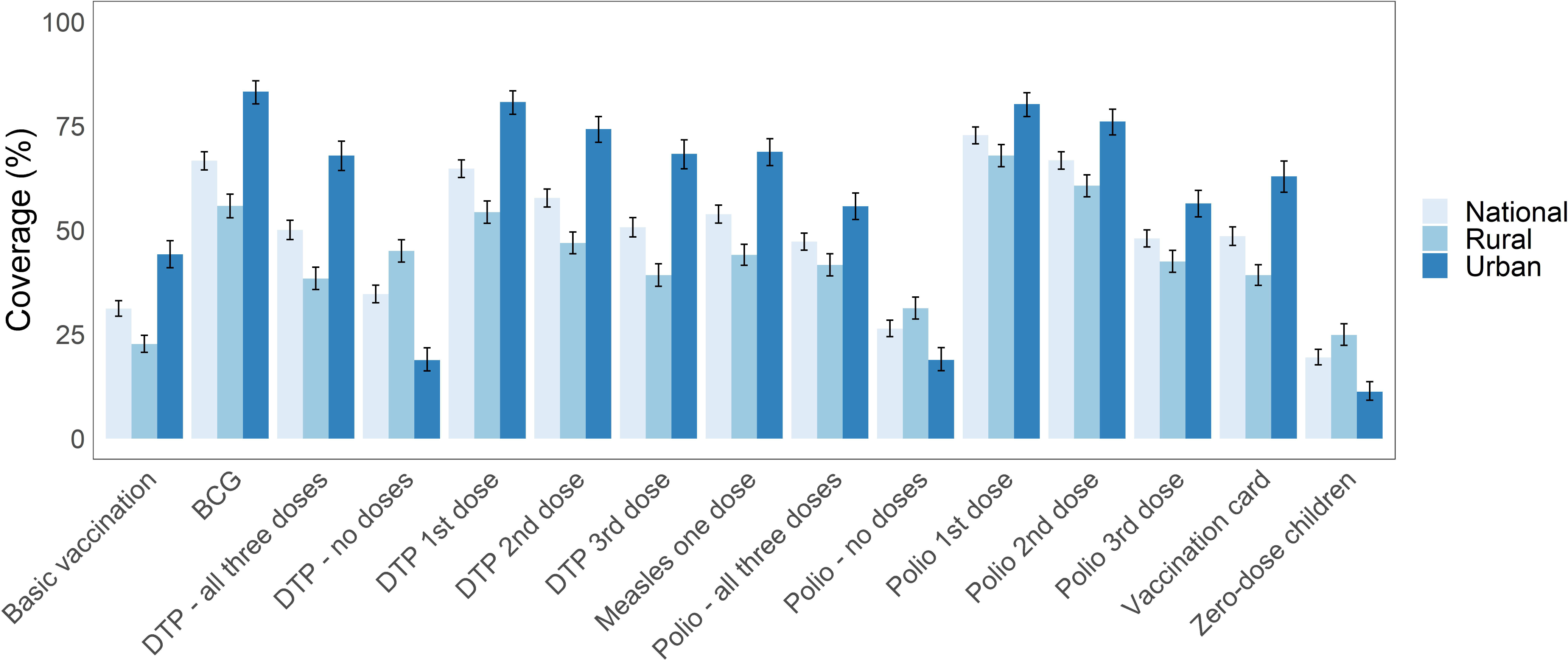
Vaccination coverage and vaccination card usage rates in Nigeria. Vaccination coverage among children aged 12-23 months in Nigeria and disaggregated by urban and rural areas of residence. Vaccination card coverage is relatively higher in urban areas in comparison to rural areas, and is associated with higher vaccination coverage. Basic vaccination includes 1-dose BCG, 3-dose DTP-HepB-Hib (diphtheria, tetanus, pertussis, hepatitis B and Haemophilus influenzae type B), 3-dose polio, and 1-dose measles vaccines.

Vaccination cards were available for 49% (46-51%) of children, among which 57% (55-60%) had received all basic vaccinations. Among the 51% (49-54%) of children without vaccination cards, only 6.4% (5.3-7.7%) of them had received all basic vaccinations. Almost one fifth of children (19% (18-21%)) had not received any of the basic vaccinations and in rural areas almost a quarter had received none. For polio vaccinations, 26% (24-28%) of children had received none of the three doses and for DTP vaccinations, 35% (33-37%) of children had received none. There was a higher proportion of zero-dose children in rural areas compared to urban areas.

### Inequity analysis

Figure 3 shows the basic vaccination coverage among children aged 12-23 months in Nigeria disaggregated by socioeconomic (household wealth, religion, ethnicity), geographic (region, place of residence), maternal (maternal age at birth, maternal education, maternal household head status), child (sex of child, birth order), and healthcare (antenatal care, birth setting) characteristics. For socioeconomic characteristics, children living in wealthier households had higher basic vaccination coverage ranging from 17% (15-20%) to 49% (45-53%) from the poorest to the richest wealth quintiles. For religion, basic vaccination coverage was higher among children of the Catholic faith at 49% (43-54%) while most children (58%) were of Islamic faith with relatively lower coverage of 23% (21-25%). Regarding ethnicity, basic vaccination coverage ranged from 12% (9-16%) to 56% (51-61%) among children of Fulani and Igbo ethnicities respectively. For geographic characteristics, 65% of children lived in rural areas but basic vaccination coverage was higher for children in urban areas at 44% (41-47%) in comparison to 23% (21-25%) in rural areas. At the regional level, basic vaccination coverage among children ranged from 20% (17-23%) in the North West region to 57% (51-62%) in the South East region (Figure 4). For maternal characteristics, children of mothers aged 35-49 years had higher basic vaccination coverage at 34% (31-39%) in comparison to 16% (12-21%) for children of younger mothers aged 15-19 years. Basic vaccination coverage increased with higher levels of maternal education, with basic vaccination coverage among children of mothers with no education, primary education, and secondary education or higher at 15% (13-17%), 33% (28-38%), and 47% (45-50%) respectively. Children living in female-headed households had relatively higher basic vaccination coverage of 39% (35-44%) in comparison to 30% (28-32%) in male-headed households. For child characteristics, basic vaccination coverage was similar among female and male children at 31% (29-33%) and 31% (29-34%) respectively, while coverage decreased by birth order with 36% (32-40%) and 23% (20-26%) among first-born and sixth-born respectively. For healthcare characteristics, children of women who had a higher number of antenatal care visits during their pregnancy had higher basic vaccination coverage at 41% (39-44%) for four or more visits and 11% (8.7-13%) for no or unknown number of visits. More than half of all women gave birth at home, and basic vaccination coverage among these children was relatively lower at 20% (18-22%) in comparison to 47% (44-49%) among children born in a public or private clinical facility.

**Figure 3.**
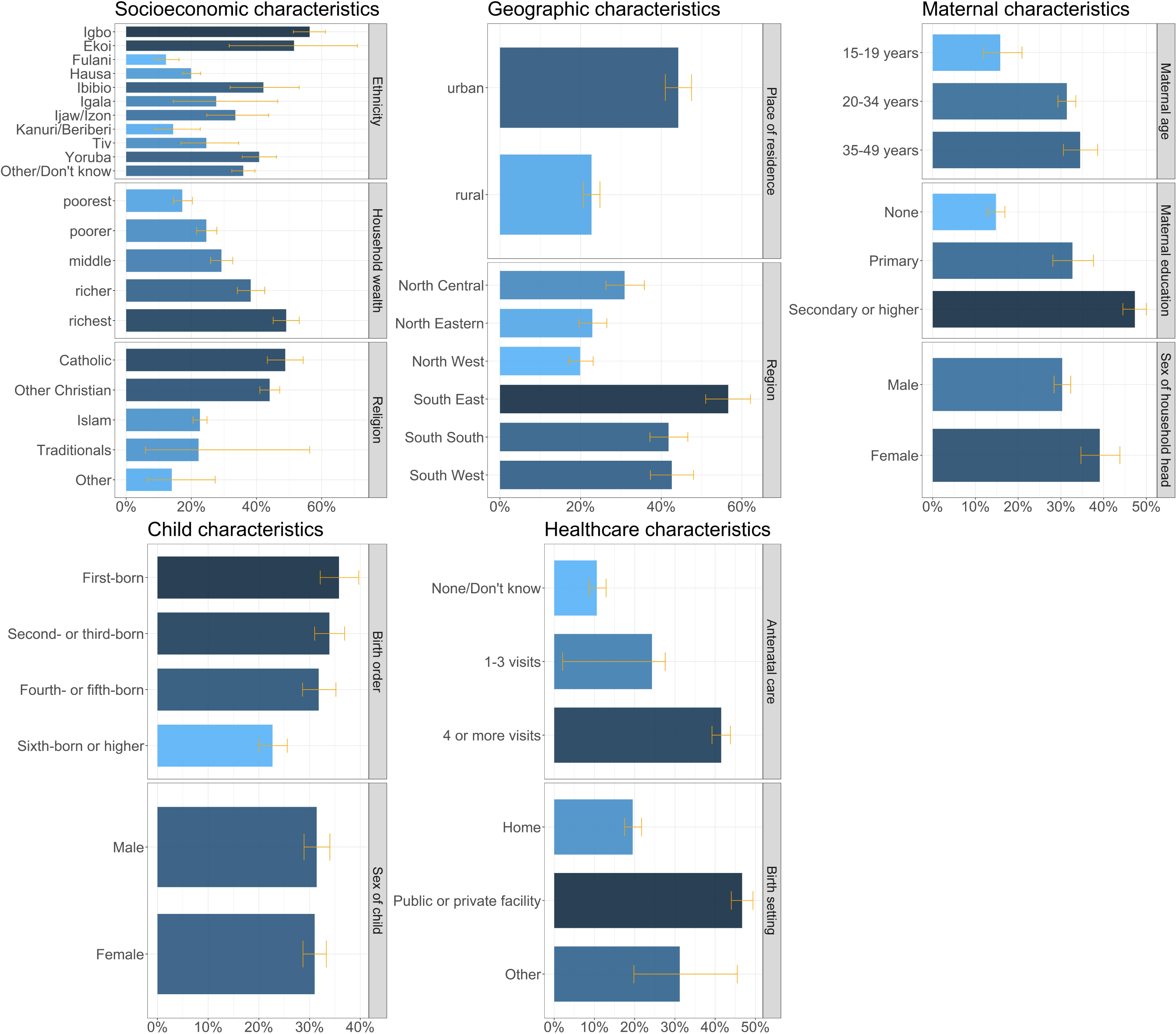
Basic vaccination coverage in Nigeria. Basic vaccination coverage among children aged 12-23 months in Nigeria by socioeconomic (household wealth, religion, ethnicity), geographic (region, place of residence), maternal (maternal age at birth, maternal education, maternal household head status), child (sex of child, birth order), and healthcare (birth setting, antenatal care) characteristics.

**Figure 4.**
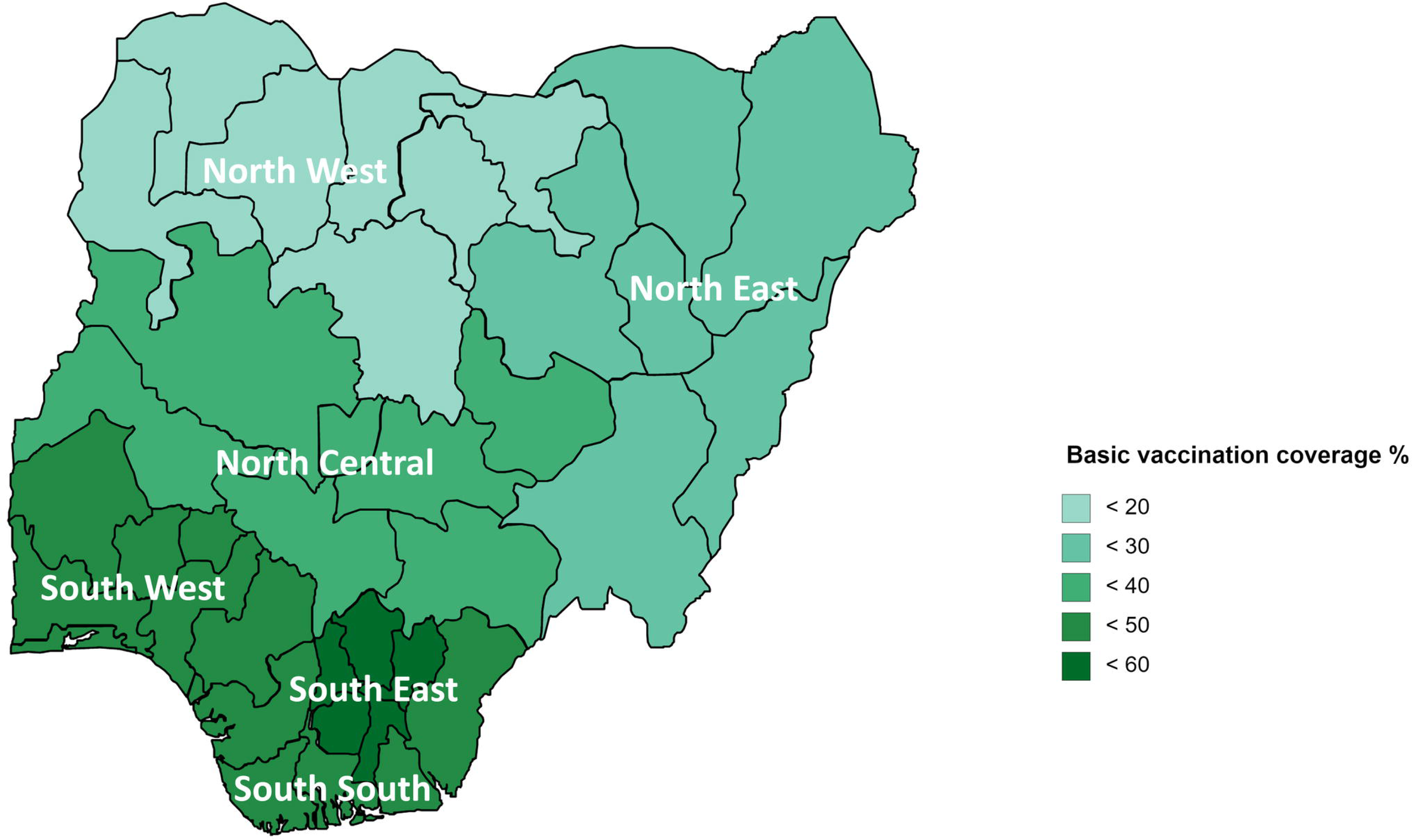
Basic vaccination coverage in Nigeria at the regional level. Basic vaccination coverage among children aged 12-23 months in Nigeria at the regional level.

Figure 5 shows the concentration curve for household wealth-related inequity in basic vaccination coverage, while Table 2 presents the inequities in vaccination card usage rates and vaccination coverage among children aged 12-23 months disaggregated by maternal education, antenatal care, and household wealth in Nigeria, based on Erreygers concentration indices. With respect to maternal education, antenatal care, and household wealth, each of these characteristics had a regressive pro-advantage distribution, with higher vaccination card usage rates and coverage among children from wealthier households, higher maternal education and more antenatal care.

**Figure 5.**
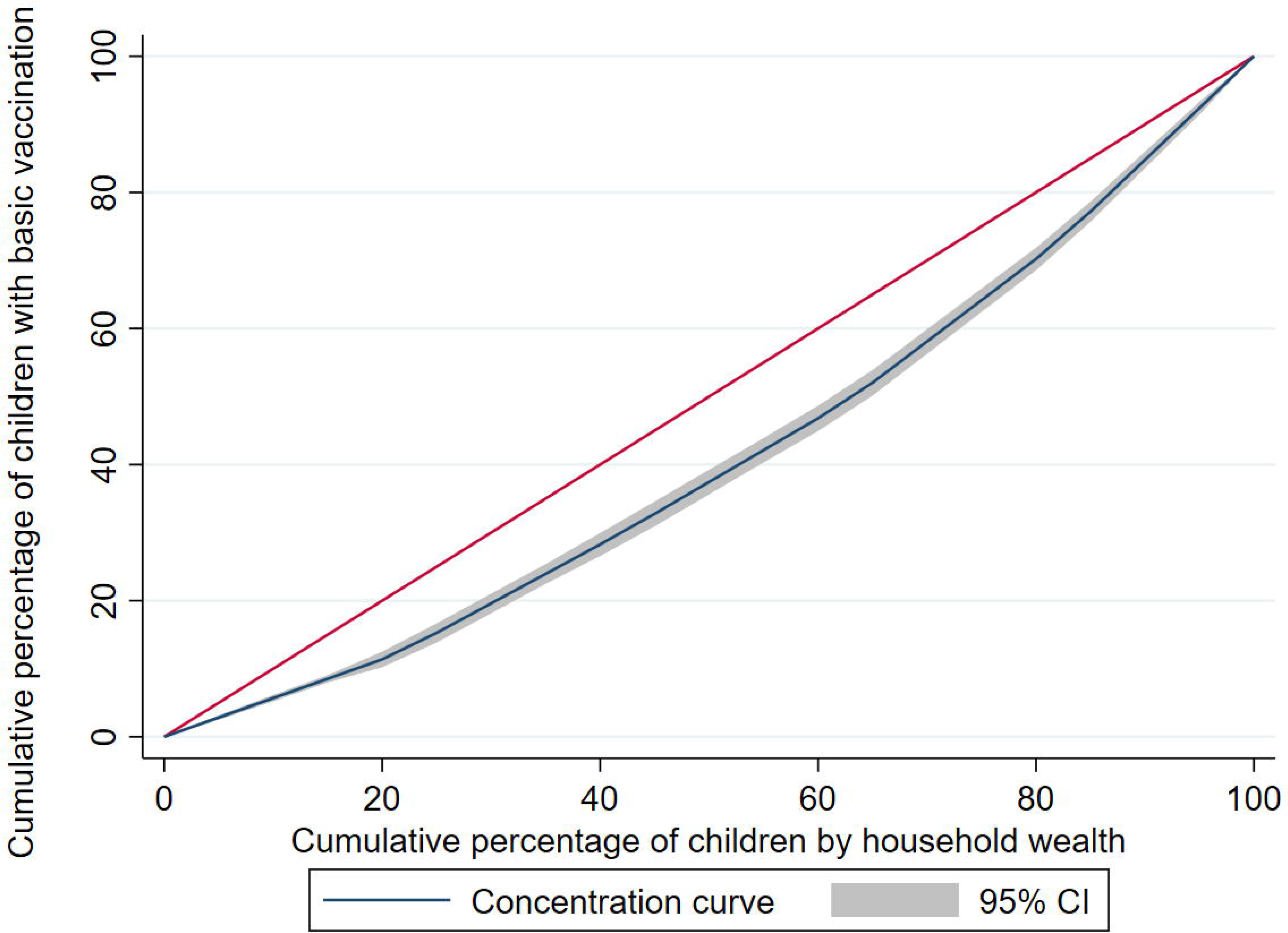
Wealth-related inequity in basic vaccination coverage in Nigeria. Concentration curve for household wealth-related inequity in basic vaccination coverage among children aged 12-23 months in Nigeria.

Inequities in vaccination card usage rates and vaccination coverage among children aged 12-23 months disaggregated by maternal education, antenatal care, and household wealth in Nigeria, based on Erreygers concentration indices (ECI).

Table 3 and Figure 6 presents the inequities in basic vaccination coverage in Nigeria among children aged 12-23 months associated with socioeconomic (household wealth, religion, ethnicity), geographic (region, place of residence), maternal (maternal age at birth, maternal education, maternal household head status), child (sex of child, birth order), and healthcare (antenatal care, birth setting) characteristics. After controlling for other background characteristics through multiple logistic regression, the AORs were significant for the associations between basic vaccination coverage and household wealth, religion, ethnicity, maternal age at birth, maternal education, antenatal care, and birth setting. This model and the AORs include the interaction between household wealth and place of residence.

Inequities in basic vaccination coverage (1-dose BCG, 3-dose DTP-HepB-Hib, 3-dose polio, and 1-dose measles) in Nigeria among children aged 12-23 months associated with socioeconomic (household wealth, religion, ethnicity), geographic (region, place of residence), maternal (maternal age at birth, maternal education, maternal household head status), child (sex of child, birth order), and healthcare (antenatal care, birth setting) characteristics. Crude and adjusted odds ratios were estimated using simple and multiple logistic regression respectively.

**Figure 6.**
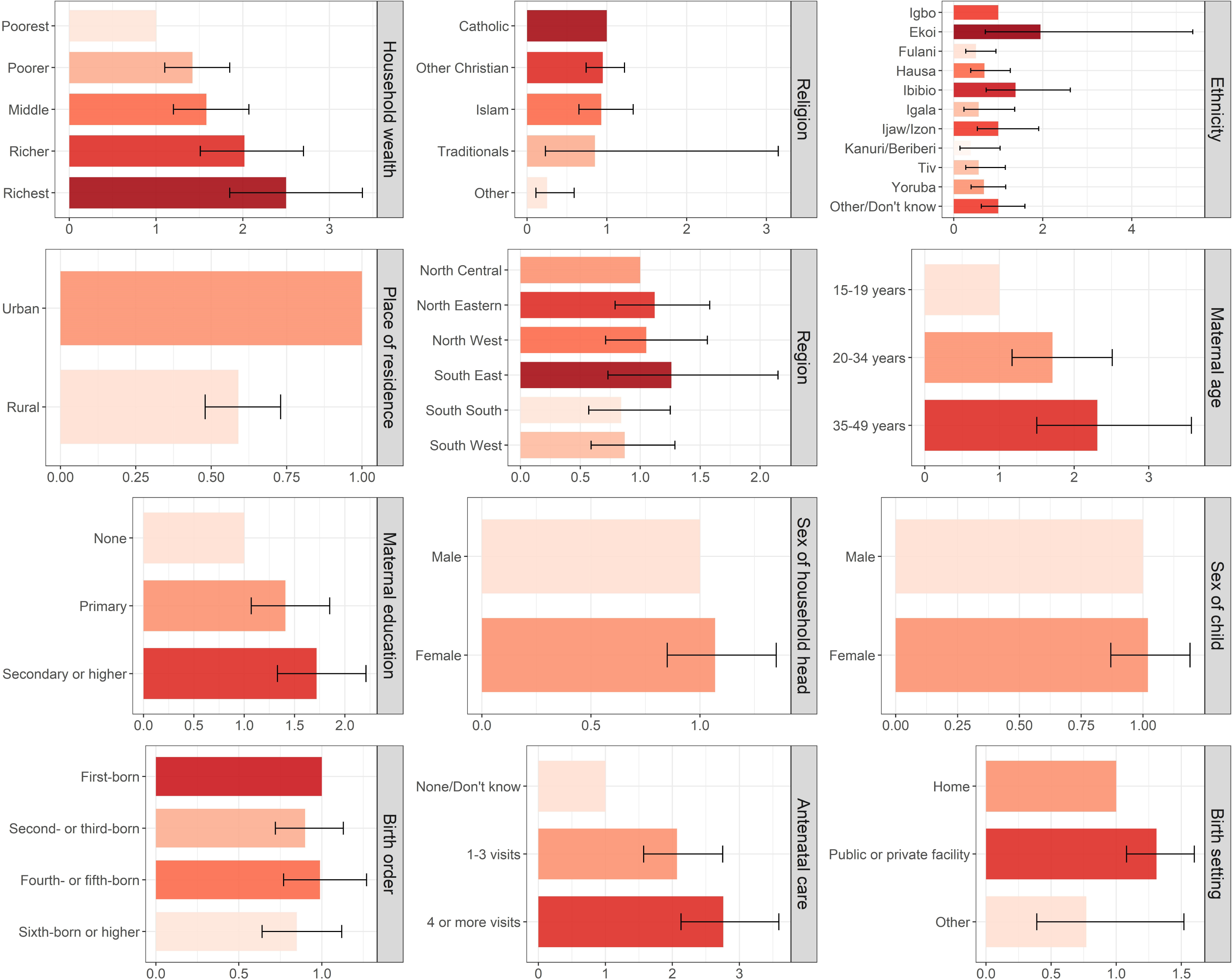
Inequities in basic vaccination coverage in Nigeria. Inequities in basic vaccination coverage among children aged 12-23 months in Nigeria associated with socioeconomic (household wealth, religion, ethnicity), geographic (region, place of residence), maternal (maternal age at birth, maternal education, maternal household head status), child (sex of child, birth order), and healthcare (birth setting, antenatal care) characteristics.

Children living in households of richest wealth quintiles had 221% higher odds (AOR: 3.21 (2.06, 5.00), p < 0.001) of receiving basic vaccinations in comparison to children of the poorest wealth quintiles. However, children living in rural areas in the richer quintile had 64% lower odds (AOR: 0.36 (0.24, 0.53), p < 0.001) and children in rural areas in the richest quintile 50% lower odds (AOR: 0.50 (0.35, 0.72), p <0.001) than children in urban areas. Children whose religion was classified as ‘other’ had 75% lower odds (AOR: 0.25 (0.10, 0.60), p < 0.002) of receiving basic vaccinations than children of Catholic faith. In comparison to children of Igbo ethnicity, children of Fulani ethnicity had 49% lower odds of receiving basic vaccinations (AOR: 0.51 (0.26, 0.97), p = 0.039). Children of mothers aged 35-49 years had 125% higher odds (AOR: 2.25 (1.46, 3.49), p < 0.001) of receiving basic vaccination than children of mothers aged 15-19 years . Children of mothers with secondary or higher education had 79% higher odds (AOR: 1.79 (1.39, 2.31), p < 0.001) of receiving basic vaccination in comparison to children of mothers with no formal education. Children of mothers who had four or more antenatal care visits had 187% higher odds (AOR: 2.87 (2.21, 3.72), p < 0.001) of receiving basic vaccinations than children of mothers who had no antenatal care. Children born in clinical facilities had 32% higher odds (AOR 1.32 (1.08, 1.61), p = 0.006) than children born in home settings of receiving basic vaccinations.

## Discussion

We conducted a systematic review to identify the social determinants of childhood immunisation in low- and middle-income countries. We selected household wealth, religion, and ethnicity for socioeconomic characteristics; region and place of residence for geographic characteristics; maternal age at birth, maternal education, and maternal household head status for maternal characteristics; sex of child and birth order for child characteristics; and antenatal care and birth setting for healthcare characteristics.

Based on the characteristics identified from the systematic review, we applied a social determinants framework to assess basic vaccination coverage (1-dose BCG, 3-dose DTP-HepB-Hib, 3-dose polio, and 1-dose measles) among children aged 12-23 months in Nigeria using the 2018 Nigeria DHS survey dataset.

The associations identified in this study between basic vaccination coverage and socioeconomic, geographic, maternal, child, and healthcare characteristics identified are supported by other studies. Basic vaccination coverage was associated with household wealth, and families in the richest quintile are more likely to live in urban areas with better access to functional private and public health facilities that provide immunisation services [24, 30]. Mothers in urban areas are more likely to use preventive healthcare services including childhood immunisation, due to their proximity to healthcare facilities in urban settings and higher educational status in comparison to mothers in rural areas with higher travel costs [31–33]. Children in the richer households of rural areas had reduced odds of basic vaccination coverage in comparison to children in the poorest households of urban areas. Children of Fulani ethnicity had lower odds of basic vaccination compared to Igbo children, and there is evidence that awareness of immunisation is low amongst Fulani mothers [34, 35]. Further, Fulani ethnic groups reside in mostly rural settings and are nomadic, limiting their access to health services including immunisation services.

We found that basic vaccination coverage was associated with maternal education. Educated mothers have better awareness and knowledge on childhood immunisation and are more able to overcome cultural barriers to vaccination [36]. Maternal age was associated with basic vaccination coverage and older mothers may have more experience with antenatal clinics and have greater awareness of immunisation services from previous children [22]. They are also more likely to have financial access to immunisation services and live in urban settings with improved access to immunisation services.

Delivery in a health facility and antenatal care were associated with basic vaccination, and to give birth in a health facility indicates that mothers have overcome barriers to accessing health services, and mothers will also receive information on childhood immunisation from healthcare workers there. Hence, utilisation of health services by mothers leads to improved immunisation status of their children [37].

At the regional level, basic vaccination among children ranged from the lowest coverage in the North West region to nearly three times higher coverage in the South East region, although residents in Northern Nigeria are more likely to have immunisation services within 5 km [38]. The large degree of autonomy of different states and the impact of ongoing conflict in parts of the country, in addition to socio-cultural reasons, can explain in part the geographical disparity in immunisation services [14]. However, regional differences in coverage were not significant in the multivariable logistic regression model.

Despite vaccination being provided at no cost to individuals, coverage was still below target levels for even the most advantaged groups and therefore we recommend a proportionate universalism approach with actions proportionate to the level of disadvantage [39, 40]. There is some evidence of payment being required for vaccination, even in public health facilities, and this may threaten vaccination uptake [32]. Both the oversight of immunisation services and public awareness of vaccinations’ no cost status should be strengthened.

Higher coverage with more antenatal care indicates greater engagement with health services and thereby providing more opportunities for vaccine education. The integration of immunisation services to nutrition programmes and paediatric outpatient departments of primary healthcare centres has been shown to improve coverage and decrease drop-out rates in South Sudan [41]. Hence, we recommend efforts to engage with families during health care visits outside of routine immunisation services and to take advantage of these missed opportunities to reach under-immunised children [42].

Our estimates of vaccination coverage in Nigeria for 2018 were lower than national estimates based on administrative reporting from health service providers, though closer to WHO and UNICEF Estimates of National Immunization Coverage (WEUNIC) (see Appendix A5) [43]. The coverage gaps have been shown to be systematically underestimated by administrative reporting in Nigeria [44]. In 80% of Nigerian states, the basic vaccination coverage was below the national target of 95% or higher coverage and below the Gavi target of 90% DTP3 coverage [45]. The below target coverage for first doses indicates challenges with access to immunisation services while the decrease in coverage for subsequent doses indicates drop-out due to insufficient knowledge on dose completion [9,46,47]. The COVID-19 pandemic has disrupted vaccination globally and coverage was lower than expected in Nigeria in 2020 [48], highlighting the urgent need for catch-up vaccination to close the immunity gaps and prevent vaccine-preventable disease outbreaks [49]. The role of conflict on these disparities is also important to understand. A literature review of conflict and vaccination inferred that conflict-affected countries had vaccination coverage below global levels and conflict affected vaccination services and human resources, including attacks on healthcare workers, which has happened in Nigeria [14, 50]. Our study inferences complement the findings in related studies in Nigeria and other countries (see Table 1).

The proportion of zero-dose children was high and raises the risk of vaccine-preventable disease outbreaks, with nearly 25% of children in rural areas not receiving a single dose of any of the basic vaccinations. Zero-dose children are also likely to lack access to health and welfare services and to suffer from multiple sources of deprivation [51, 52]. More than 50% of children do not have vaccination cards and nearly 60% of children do not have birth registrations [53] - improving uptake of vaccination cards and birth registrations would help in part to address the barriers for vaccine access for all children, including zero-dose children.

Our study has limitations, and we cannot estimate causal-effect relationships nor temporal inferences due to the cross-sectional study design of 2018 Nigeria DHS. Our study has similar biases that are associated with DHS surveys, including recall bias, measurement bias, and social desirability bias which tend to overestimate vaccination coverage. In particular, only 49% of children had a vaccination card and when a vaccination card was not available for a child, their mother was asked to recall their vaccinations, which may lead to an overestimation of coverage.

For future work, we recommend qualitative research to understand the barriers and enablers of childhood vaccination and their associations with socioeconomic, geographic, maternal, child, and healthcare characteristics which would be valuable to adapt vaccination programmes to improve coverage equitably in Nigeria.

## Conclusions

We identified the inequities in basic vaccination coverage by socioeconomic, geographic, maternal, child, and healthcare characteristics among children aged 12-23 months in Nigeria using a social determinants of health perspective. In conclusion, we infer that inequities in basic vaccination were associated with lower coverage among children living in poorer households, belonging to Fulani ethnicity, born in home settings in Nigeria, with younger mothers at birth, with mothers with no formal education and with mothers who had no antenatal care visits. We recommend a proportionate universalism approach with targeted vaccination programmes proportionate to the level of disadvantage for addressing the immunisation barriers faced by these underserved subpopulations. This will improve coverage and reduce inequities in childhood immunisation associated with socioeconomic, geographic, maternal, child, and healthcare characteristics in Nigeria.

## Abbreviations

AOR: Adjusted odds ratio
BCG: Bacille Calmette-Guérin vaccine
DHS: Demographic Health Survey
DTP: Diphtheria, tetanus and pertussis containing vaccine
DTP-HepB-Hib: Diphtheria, tetanus, pertussis, hepatitis B and Haemophilus influenzae type B ECI Erreygers concentration indices
EPI: Expanded Programme of Immunization
Gavi: Global Alliance for Vaccines and Immunisations
LGA: Local Government Area
LMIC: Low- and Middle-Income Countries
PRISMA: Preferred Reporting Items for Systematic Reviews and Meta-Analyses
SDG: Sustainable Development Goals UNICEF United Nations Children’s Fund WHO World Health Organisation
WUENIC: WHO and UNICEF estimates of national immunization coverage

## Declarations

### (a) Ethics Approval and Consent to Participate

This study was approved by the ethics committee (Reference: 21600) of the London School of Hygiene & Tropical Medicine. For DHS surveys in general, the procedures and questionnaires are approved by the ICF Institutional Review Board (IRB), and the country-specific DHS survey protocols are reviewed by the ICF IRB and an IRB in the host country. Before each interview is conducted for the DHS survey, an informed consent statement is read to the respondent, who may accept or decline to participate. Further information on informed voluntary participation in the DHS survey is available here: https://dhsprogram.com/Methodology/Protecting-the-Privacy-of-DHS-Survey-Respondents.cfm All methods in this study were carried out following the relevant rules and regulations.

### (b) Consent for Publication

Not applicable.

### (c) Availability of data and materials

The analysis code is publicly accessible on GitHub at https://github.com/svwilliams122/vaccine-equity-Nigeria. The 2018 Nigeria DHS data set is publicly available and accessible upon registration for legitimate research purposes on the DHS website at https://dhsprogram.com/methodology/survey/survey-display-528.cfm. To download DHS datasets, researchers must register as a DHS data user at https://dhsprogram.com/data/new-user-registration.cfm

### (f) Acknowledgment

We thank the DHS Program for access to the 2018 Nigeria Demographic and Health Survey dataset.

### (g) Author’s Contribution

SVW and KA conceptualised the study. SVW undertook the analysis and wrote the initial draft. TA provided contextual expertise of the national immunisation programme in Nigeria to explain the findings. SVW, TA, and KA contributed to the analysis, interpretation of results, and the final drafting of the manuscript. All authors have read and approved the final manuscript.

## Data Availability

All data produced are available online at https://dhsprogram.com/data/

