## Appendices for "Childhood vaccination coverage by socioeconomic, geographic, maternal, child, and healthcare characteristics in Nigeria: equity impact analysis"

**Appendix**

**Table A1. Routine vaccination schedule of under 1-year-old children in Nigeria.**

| Vaccine | Schedule |
| --- | --- |
| BCG (Bacille Calmette-Guérin) | Birth |
| Hepatitis B (birth dose) | Birth |
| Oral polio | Birth  6, 10, 14 weeks |
| DTP-HepB-Hib (diphtheria, tetanus, pertussis, hepatitis B and Haemophilus influenzae type B) | 6, 10, 14 weeks |
| Pneumococcal conjugate | 6, 10, 14 weeks |
| Inactivated polio | 14 weeks |
| Measles first dose | 9 months |
| Meningitis A conjugate | 9 months |
| Yellow fever | 9 months |

**Figure A2. PRISMA flowchart.** The Preferred Reporting Items for Systematic Reviews and Meta Analyses (PRISMA) flow diagram of articles’ identification, screening, eligibility, and inclusion in the systematic review is illustrated.

**Screening**

**Included**

**Eligibility**

**Identification**

Records identified through database searching MEDLINE

(n = 155)

Records excluded

(n = 2)

Reason: duplication

Records screened by title and abstract

(n = 158)

Records excluded

(n = 103)

Reasons: Irrelevant or fell into exclusion categories

Full-text articles assessed for eligibility

(n = 55)

Studies included in qualitative synthesis

(n = 49)

Full-text articles excluded

(n = 6)

Reasons:

not relevant (n = 3)

not focused on LMICs (n = 2)

data analysed too old to be relevant (n = 1)

Additional records identified through other sources

(n = 5)

**Table A3. Vaccination coverage and vaccination card usage rates in Nigeria.** Vaccination card usage rates and vaccination coverage among children aged 12-23 months in Nigeria, at the national level and disaggregated by urban and rural areas of residence.

|  | National coverage  (% and 95% CI)  n = 6059 | Urban coverage  (% and 95% CI)  n1 = 2100 | Rural coverage  (% and 95% CI)  n2 = 3959 |
| --- | --- | --- | --- |
| Vaccination card | 48.6  (46.3, 50.8) | 62.9  (59.1, 66.6) | 39.2  (36.7, 41.7) |
| Vaccine | | | |
| BCG | 66.7  (64.4, 68.8) | 83.3  (80.3, 85.9) | 55.8  (53.0, 58.7) |
| Polio 1^st^ dose | 72.8  (70.7, 74.8) | 80.3  (77.3, 83.0) | 67.9  (65.2, 70.6) |
| Polio 2^nd^ dose | 66.8  (64.7, 68.8) | 76.1  (72.9, 79.0) | 60.7  (58.0, 63.3) |
| Polio 3^rd^ dose | 48.0  (46.0, 50.1) | 56.4  (53.2, 59.6) | 42.5  (39.9, 45.2) |
| Polio – all three doses | 47.2  (45.2, 49.3) | 55.8  (52.6, 58.9) | 41.7  (39.0, 44.3) |
| No doses of polio | 26.4  (24.4, 28.4) | 18.9  (16.3, 21.8) | 31.7  (28.7, 34.0) |
| DTP 1^st^ dose | 64.8  (62.6, 66.9) | 80.8  (77.8, 83.5) | 54.3  (51.7, 57.0) |
| DTP 2^nd^ dose | 57.8  (55.6, 59.9) | 74.3  (71.1, 77.3) | 47.0  (44.4, 49.6) |
| DTP 3^rd^ dose | 50.7  (48.4, 53.0) | 68.3  (64.7, 71.7) | 39.2  (36.5, 41.9) |
| DTP – all three doses | 50.1  (47.8, 52.4) | 67.9  (64.3, 71.3) | 38.4  (35.8, 41.1) |
| No doses of DTP | 34.7  (32.6, 36.8) | 18.8  (16.2, 21.8) | 45.0  (42.4, 47.7) |
| Measles one dose | 53.9  (51.7, 56.0) | 68.8  (65.5, 71.9) | 44.1  (41.6, 46.6) |
| Basic vaccinations (all) | 31.2  (29.4, 33.1) | 44.2  (41.0, 47.5) | 22.7  (20.7, 24.8) |
| No basic vaccination doses | 19.5  (17.7, 21.4) | 11.2  (9.2, 13.7) | 24.9  (22.4, 27.6) |

**Table A4. Interaction effects.**

(A) Interaction terms between place of residence and household wealth
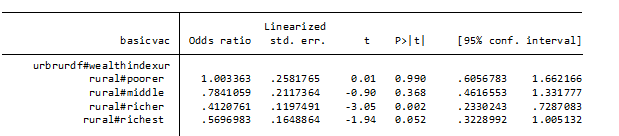

(B) AORs of basic vaccination coverage in Nigeria by household wealth in rural areas in comparison to urban areas

| Characteristics | Adjusted odds ratio | 95% confidence interval | p value |
| --- | --- | --- | --- |
| Rural – poorest | 0.88 | 0.56 – 1.37 | 0.560 |
| Rural – poorer | 0.88 | 0.60 – 1.28 | 0.499 |
| Rural – middle | 0.69 | 0.47 – 1.00 | 0.049 |
| Rural – richer | 0.36 | 0.24 – 0.53 | <0.001 |
| Rural – richest | 0.50 | 0.35 – 0.72 | <0.001 |

**Table A5. Vaccine coverage.** Vaccine coverage estimates for Nigeria in 2018 based on DHS, WUENIC (WHO-UNICEF), administrative, and official country sources.

| Source | Vaccine coverage (%) | | | |
| --- | --- | --- | --- | --- |
|  | BCG | Measles | DTP 3 | Polio 3 |
| DHS | 67 | 54 | 50 | 47 |
| WUENIC | 67 | 54 | 56 | 56 |
| Administrative – provider estimates | 90 | 87 | 95 | 95 |
| Official country estimates | 75 | 63 | 58 | 58 |
